# The association between severity and aetiology of chronic liver disease and seasonal influenza vaccination uptake in adults: a retrospective cohort study using English primary care data (2019–2024)

**DOI:** 10.64898/2026.04.08.26350434

**Authors:** Ilsa L. Haeusler, David Etoori, Colin N. J. Campbell, Suzanna L. R. McDonald, Jamie Lopez Bernal, Sandra Mounier-Jack, Ben Kasstan-Dabush, Helen I. McDonald, Edward P. K. Parker, Anne M. Suffel

**Affiliations:** Department of Infectious Disease Epidemiology and International Health, London School of Hygiene and Tropical Medicine, London, UK; NIHR Health Protection Research Unit in Immunisation at the London School of Hygiene and Tropical Medicine; Public Health Programmes Directorate, UK Health Security Agency; Department of Infectious Disease Epidemiology, Imperial College London, London, UK; Department of Global Health and Development, London School of Hygiene & Tropical Medicine, London, UK; Global Health Policy Unit, University of Edinburgh, Edinburgh, UK; Department of Life Sciences, University of Bath, Bath, UK; Department of Non-communicable Disease Epidemiology, London School of Hygiene and Tropical Medicine, London, UK

**Keywords:** influenza, vaccine, uptake, chronic liver disease, electronic health records

## Abstract

**Objective:** To examine the relationship between chronic liver disease (CLD) severity and aetiology, and influenza vaccine uptake in England.

**Methods:** A retrospective cohort study of adults using Clinical Practice Research Datalink Aurum was conducted for five seasons (2019/20–2023/24). Poisson regression was used to estimate uptake by CLD severity (low, moderate, or severe) and aetiology (alcohol-related, viral-related, and diagnoses in the ‘Green Book’ guidelines).

**Results:** Among individuals with CLD who were additionally age-eligible for vaccination, uptake was 71.1–79.7% versus 30.9–40.5% in those not age-eligible. Among individuals below age eligibility without other comorbidities, severity was associated with higher uptake (incidence rate ratio [IRR] moderate 1.89 [95% CI 1.80–1.99]; severe 2.12 [2.01–2.23] in 2023/24); there was no effect in those with at least one additional comorbidity. Alcohol- and viral-related aetiology were also associated with increased uptake in those not additionally age-eligible. Among individuals meeting age eligibility without additional comorbidities, severity was associated with reduced uptake (moderate 0.85 [0.78–0.93]; severe 0.80 [0.76–0.85]), with attenuation in those with additional comorbidities (moderate 0.97 [0.92–1.02]; severe 0.91 [0.89–0.94]).

**Conclusions:** CLD severity and aetiology were important determinants of uptake in the absence of additional indications for influenza vaccination.

## Introduction

Influenza is a disease of major global public health significance.^1^ In England, the influenza vaccination programme aims to directly protect high-risk individuals by offering free annual vaccination to adults aged 65 years and older, those aged six months to under 65 with certain underlying health conditions (‘clinical risk groups’), and pregnant individuals (alongside other groups to also reduce community transmission).^2^ General practitioners (GPs) and pharmacies (and maternity services for pregnant women) are commissioned to deliver the seasonal influenza programme and are reimbursed per vaccine delivered.^3,4^

England has adopted a coverage ambition of at least 75% among adults aged 65 years and older, aligned with WHO European Council recommendations.^5^ For clinical risk groups aged six months to under 65 years, coverage ambition was at least 55% in 2019/20 (temporarily increased to 75% during 2020/21 due to the COVID-19 pandemic).^6^ During 2024/25, coverage among adults aged 65 years and older was 74.9% compared to 40.0% among those aged six months to 65 years in clinical risk groups.^7,8^ Individuals with chronic liver disease (CLD) consistently have amongst the lowest coverage of all clinical risk groups, most recently at 33.8% among those aged six months to 65 years.^8^

CLD causes immune dysregulation, placing those affected at substantially increased risk of influenza-related complications.^9^ During the 2010/11 season in England, individuals with CLD had the highest age-adjusted mortality rate among clinical risk groups.^2^ In the 2013/14 Northern Hemisphere season, people with CLD had almost twice the odds of hospitalisation with influenza compared to those without underlying comorbidities.^10^ Mortality rises with increasing severity of CLD following a broad range of infections,^11^ and with alcohol-related CLD following COVID-19.^12^ The same likely applies to influenza, given infection can trigger hepatic decompensation.^13^

Individuals within CLD are heterogeneous in terms of severity of disease, specific diagnoses and underlying aetiology. It is important that all individuals with CLD benefit from equitable access to vaccination, particularly those at highest risk of severe complications. We therefore aimed to investigate the association of CLD severity and aetiology with seasonal influenza vaccine uptake among primary care-registered adults in England.

## Methods

### Data sources

We used data from Clinical Practice Research Datalink (CPRD) Aurum^14^ – a research service providing longitudinal, anonymised, routinely collected data from participating primary care practices. Data include demographic details, diagnoses, and procedures, including vaccinations. We linked small area data to obtain Index of Multiple Deprivation (IMD) values based on individual household postcode.^15^ When individual-level linkage could not be performed, practice-level IMD was used.

As of June 2024, CPRD Aurum included 16.2 million individuals meeting quality standards registered at currently-contributing practices, representing approximately 24.2% of the UK population.^14^ CPRD Aurum is considered broadly representative of the English population with respect to age, sex, and socioeconomic status.^16,17^

### Study design and population

Separate retrospective cohorts were defined for each influenza season (1st September to 28th February 2019/20–2023/24). Each season included adults aged 18–115 years with a recorded CLD diagnosis. Individuals were required to be registered for at least one year prior to cohort entry to ensure sufficient time for medical records to be inputted or transferred from a previous practice. Records were excluded if they did not meet CPRD’s data quality criteria^18^ and additional data processing checks (Appendix 1).

### Exposures

CLD codelists were developed by medically qualified investigators according to best practice guidelines.^19^ This included search terms for any previous clinical diagnosis of, or clinical review specific to, CLD including cirrhosis, steatosis, oesophageal varices, biliary atresia and chronic hepatitis.^20^ The primary exposure was CLD severity. There is no overall severity classification across all CLD diagnoses in clinical practice. Classification of CLD severity was therefore based on association of diagnoses with risk of severe infection and mortality: steatosis (low severity); liver transplant, cirrhosis, and varices (severe); all other diagnoses including viral hepatitis, chronic hepatitis, and biliary atresia (moderate severity).^21,22^ If an individual had more than one relevant code, the most severe diagnosis was used. Secondary exposures were CLD aetiology (Appendix 1). These were alcohol-related (classified as not alcohol-related, uncertain if alcohol-related, and alcohol-related) and viral-related (not viral-related, uncertain if viral-related, and viral-related). These were chosen because they represent two of the most common underlying causes of CLD in the UK.^23^ The final exposure was whether the diagnosis was explicitly listed as an example CLD diagnosis in the Green Book guidelines (cirrhosis, biliary atresia, chronic hepatitis diagnoses were classified as listed, all other diagnoses were not).^2^

### Outcome

The outcome was influenza vaccine administration as a binary variable. Follow-up began on 1st September for each season and continued until the earliest of: vaccination, death (earliest date recorded in CPRD or ONS mortality data), end of patient’s registration, last data collection date from the practice, or 28 February the following year. Vaccine administration was identified using influenza vaccine-related codes (according to methods previously validated for ascertaining childhood vaccination status in CPRD Aurum; Appendix 1).^24^

### Covariates

Available covariates were age, sex, region, ethnicity, IMD as a measure of socioeconomic deprivation, and other comorbidities which qualify individuals for seasonal influenza vaccination^2^ (Appendix 1 for variable definitions). The only clinical risk group not available was pregnancy. A binary variable was created (‘any other clinical risk comorbidity’) to indicate whether an individual had at least one additional vaccine-eligible comorbidity. If a qualifying comorbidity was not recorded, it was assumed not present.

### Statistical analysis

Analyses were conducted in R (v4.5.0). All analyses were stratified into cohorts which were eligible for influenza vaccination due to CLD (not additionally eligible due to age) and those which were additionally eligible due to age (≥65 years in 2019/20 and 2023/24, and ≥50 years in 2020/21, 2021/22, and 2022/23).

Incidence rate ratios (IRRs) of vaccine uptake by CLD severity and aetiology were estimated using Poisson regression including log person-time at risk as an offset with robust standard errors. For each exposure, season, and cohort, we explored associations adjusting sequentially for potential confounders. Model 1 included the exposure and age group (all individuals); model 2 added clinical risk comorbidity (all individuals); model 3 added sex, geographical region and IMD (all individuals), and model 4 (fully-adjusted) added ethnicity (thus restricted to those with complete ethnicity). To assess the impact of potential selection bias from missing ethnicity data, a sensitivity analysis was conducted by rebuilding model 3 using a dataset limited to those with complete ethnicity. Individuals with indeterminate gender were excluded due to very small numbers.

For the most recent season (2023/24), potential variation in the association between CLD severity and vaccine uptake was assessed by performing analyses stratified by clinical risk comorbidity, ethnicity and IMD using the fully adjusted model (model 4).

Our findings are reported in accordance with RECORD guidelines (Appendix 2).^25^

## Results

The number of individuals with CLD in each season from 2019/20–2023/24 was 182,174, 203,666, 228,180, 251,734, and 277,470, respectively (Appendix 3). Median follow-up was 100 days (IQR 41.0–180.0), 67.0 (IQR 32.0–180.0), 68.0 (IQR 36.0–180.0), 68.0 (IQR 37.0–180.0), and 74.0 (IQR 34.0–180.0). The number of individuals who exited follow-up without vaccination before the end of each season ranged from 5,490 (2.2%; 2022/23) to 7,812 (2.8%; 2023/24).

Demographic characteristics were broadly similar between seasons (Table 1). In the cohorts eligible for vaccination due to CLD (hereafter ‘younger’ cohorts), the proportion of males was 54.7–60.7%, compared with 46.3–50.2% in cohorts eligible due to both CLD and age (‘older cohorts’). Whilst White ethnicity was most common in both younger (56.8–71.5%) and older cohorts (71.7–85.0%), younger cohorts included a larger proportion of individuals from each ethnic minority group. In younger cohorts, there was a higher proportion of individuals living in more socioeconomically deprived areas (25.7–28.3% most deprived; 13.1–15.3% least deprived) while the distribution was more even in older cohorts. The presence of at least one other clinical risk comorbidity was more common in older cohorts (63.7–73.7%) compared to younger cohorts (37.9–49.4%), most commonly diabetes (Appendix 4). Although most individuals had low severity CLD in both younger (80.6–85.8%; Appendix 5) and older cohorts (76.3–82.5%; Appendix 6), the proportion with severe disease was higher in the older cohorts.

**Table 1:**
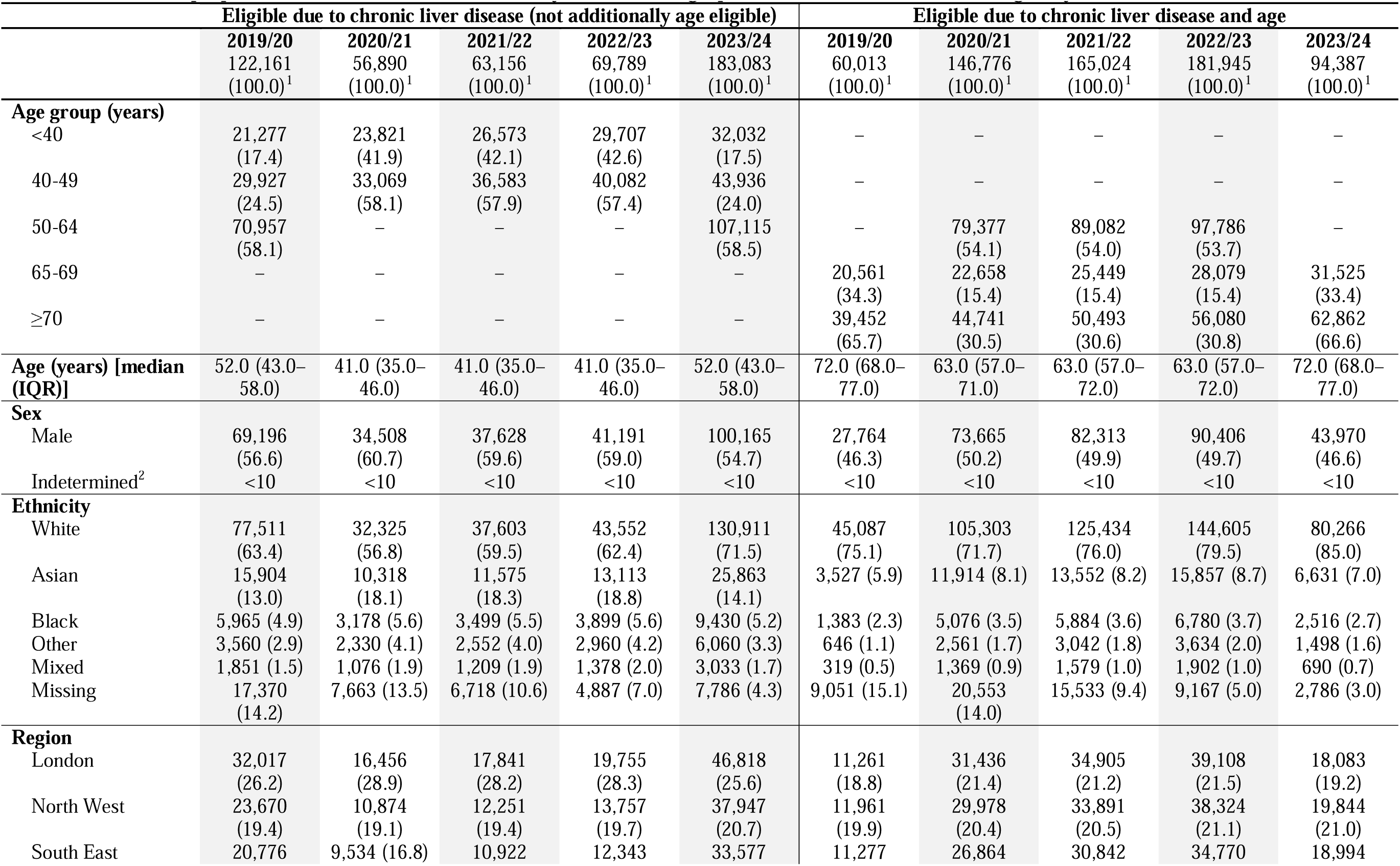
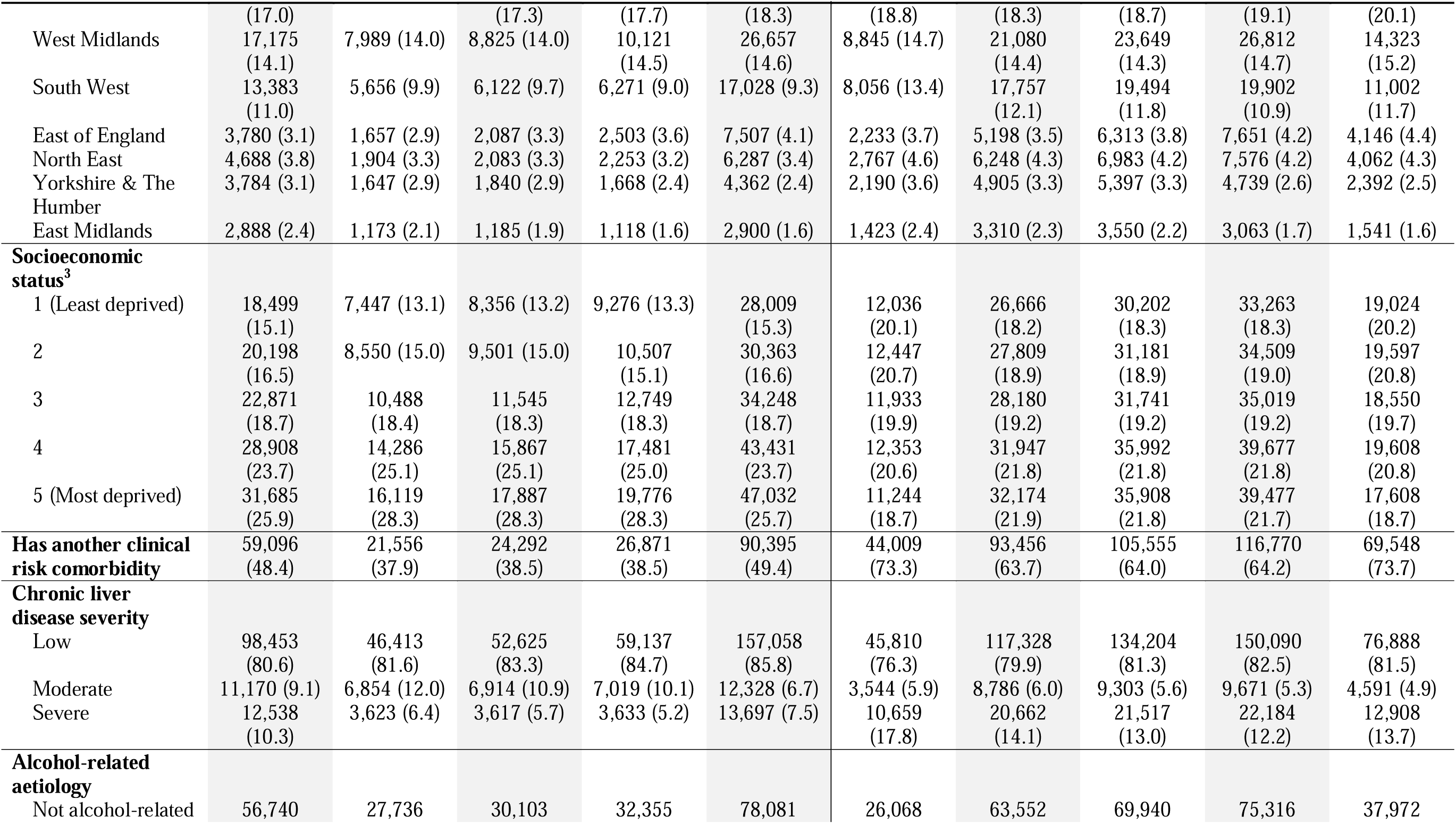
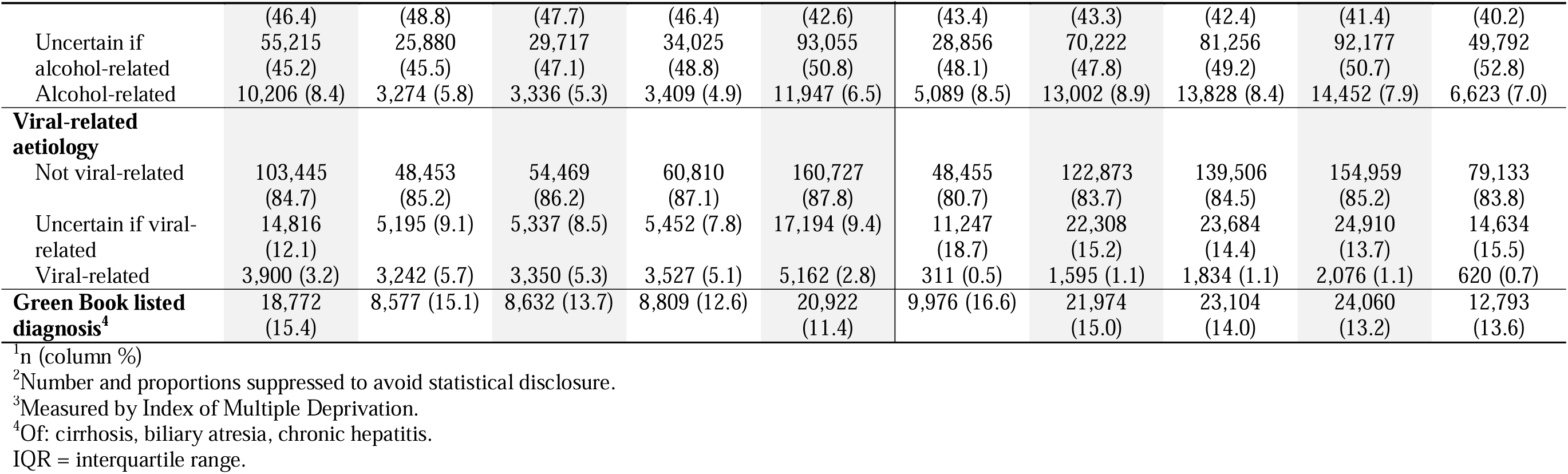
Number and proportion of individuals in each cohort, by baseline demographic characteristics and vaccine eligibility.

Vaccine uptake in the younger cohorts ranged between 30.9% (2022/23) and 40.5% (2019/20; Appendix 7), with uptake increasing by severity (44.6–53.6% in severe CLD; Figure 1). In older cohorts, uptake ranged between 71.1% (2022/23) and 79.7% (2023/24; Appendix 8) with no consistent trend by CLD severity. Overall, there was a strong age-based gradient in uptake (Appendices 7 and 8).

**Figure 1:**
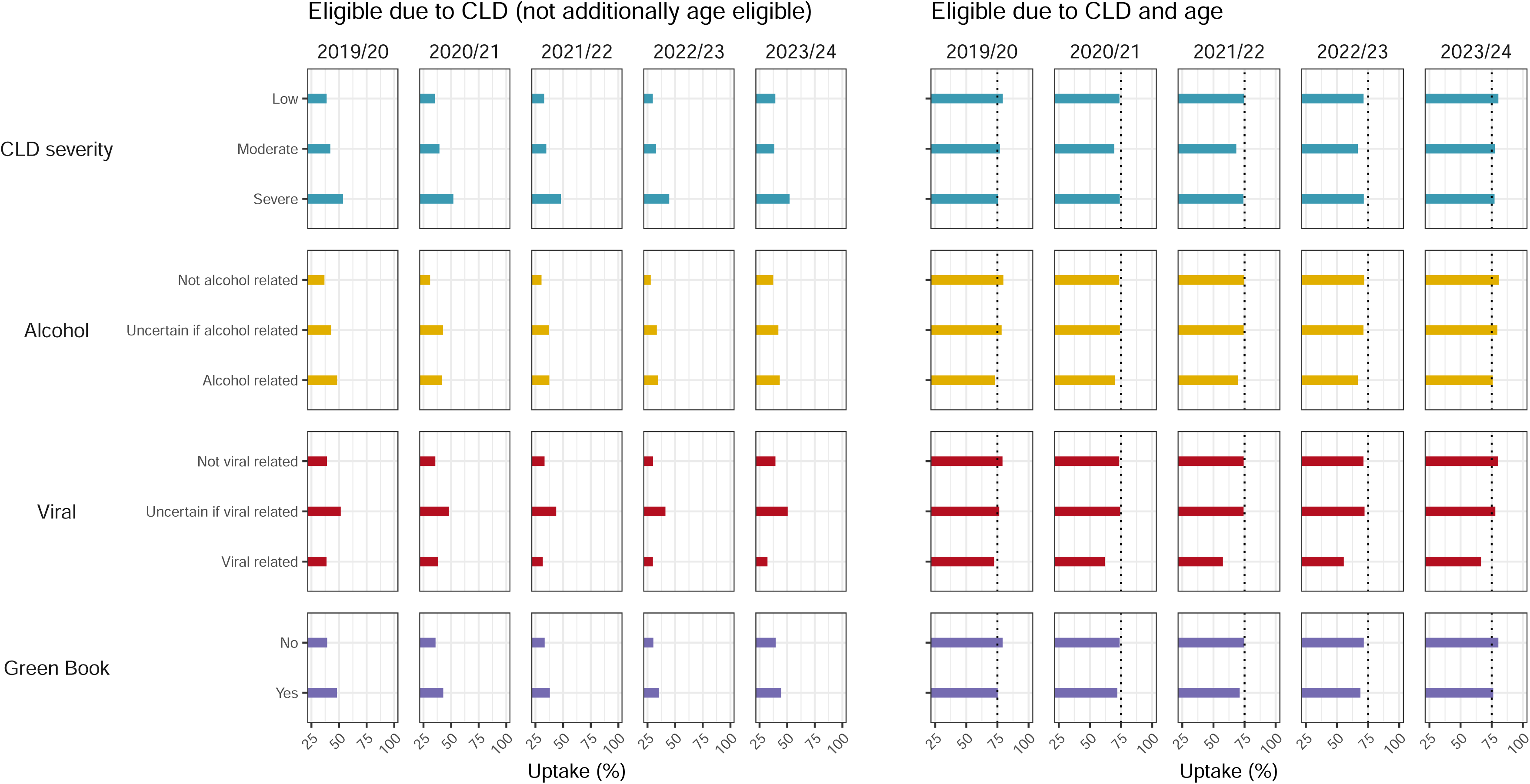
Proportion of individuals that received influenza vaccination per season, by exposure (chronic liver disease severity, alcohol-aetiology, viral-aetiology, and Green Book listed diagnosis) and influenza vaccination eligibility. Vertical dotted line: influenza vaccine coverage ambition for the age-based programme during each influenza season in England. Influenza seasons: 1st September to 28th February inclusive. In 2019/20 and 2023/24 individuals aged ≥65 years were eligible for influenza vaccination (irrespective of additional clinical risk comorbidities). In 2020/21, 2021/22, and 2022/23 age eligibility was widened to ≥50 years due to the COVID-19 pandemic.

In younger cohorts, moderate and severe disease were associated with higher fully-adjusted IRRs of vaccine uptake compared to low severity disease (IRR moderate 1.27 [95% CI 1.22–1.32] to 1.45 [95% CI 1.40–1.50]; IRR severe 1.22 [95% CI 1.18–1.27] to 1.52 [95% CI 1.43–1.61]; Figure 2 and Appendix 9). In older cohorts, increasing severity was associated with a modest reduction in vaccine uptake (IRR moderate 0.97 [95% CI 0.93–1.00] to 0.92 [95% CI 0.89–0.95]; IRR severe 0.95 [95% CI 0.93–0.97] to 0.89 [95% CI 0.86–0.91]). There were no important differences in model estimates when excluding individuals with missing ethnicity data (Appendix 9).

**Figure 2:**
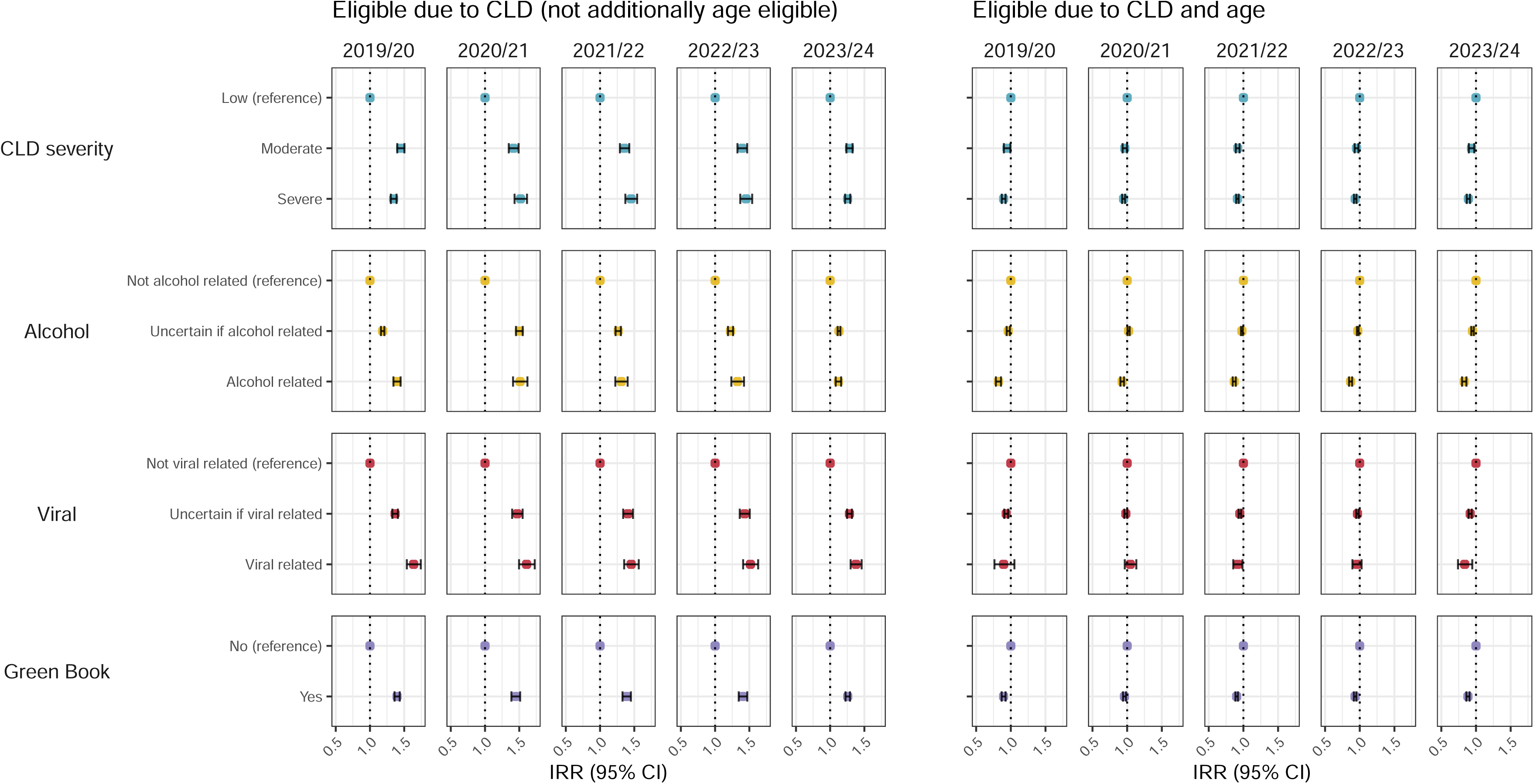
Incidence rate ratios of influenza vaccine uptake per season, by exposure (chronic liver disease severity, alcohol-aetiology, viral-aetiology, and Green Book listed diagnosis) and influenza vaccination eligibility. Calculated using Poisson regression with robust standard errors. Estimates were derived from fully-adjusted models (model 4), adjusted for age group, other clinical risk comorbidity, sex, socioeconomic status, geographical region, and ethnicity. In 2019/20 and 2023/24 individuals aged ≥65 years were eligible for influenza vaccination (irrespective of additional clinical risk comorbidities). In 2020/21, 2021/22, and 2022/23 age eligibility was widened to ≥50 years due to the COVID-19 pandemic. CLD = chronic liver disease; CI = confidence interval; IRR = incidence rate ratio

In stratified analyses, among younger individuals without additional clinical risk comorbidities during 2023/24, uptake was 23.9% and strongly associated with severity (IRR moderate 1.89 [95% CI 1.80–1.99]; severe 2.12 [95% CI 2.01–2.23]), whereas in those with at least one additional clinical risk comorbidity, uptake was 56.7% and the effect of CLD severity was minimal (moderate 0.99 [95% CI 0.94–1.04]; severe 1.02 [95% CI 0.98–1.06]; Appendix 10). Among older individuals without additional comorbidities, uptake was 77.2% in 2023/24 and severity was associated with a reduction in vaccine uptake (moderate 0.85 [95% CI 0.78–0.93]; severe 0.80 [95% CI 0.76–0.85]), with higher uptake (80.6%) and attenuation of the effect of severity in those with additional comorbidities (moderate 0.97 [95% CI 0.92–1.02]; severe 0.91 [95% CI 0.89–0.94]).

Among younger individuals, larger effect sizes for moderate and severe disease versus low severity disease were generally observed among individuals from ethnic minorities compared to White individuals (Appendix 11). These estimates were relatively imprecise with some overlapping confidence intervals. Among older individuals, there was no clear modifying effect of ethnicity. Socioeconomic status modified the association in younger individuals, whereby increasing socioeconomic deprivation was associated with attenuation of the effect of severity on uptake (Appendix 12). This trend was not observed among older individuals.

Alcohol-related CLD accounted for 5.3–8.4% of younger and 7.0–8.9% of older cohorts (Table 1). Among younger cohorts, uptake was lower in those with non-alcohol-related CLD (27.9–37.4%) compared to alcohol-related aetiology (34.5–48.3%, IRR 1.12 [95% CI 1.08–1.16] to 1.51 [95% CI 1.41–1.62]) and uncertain alcohol aetiology (33.4–42.9%, IRR 1.13 [95% CI 1.11–1.15] to 1.50 [95% CI 1.45–1.55]; Figures 1 and 2, Appendices 7 and 13). Among older cohorts, the association reversed with uptake lowest in those with alcohol-related aetiology (66.7–75.8% compared to 71.8–80.6% non-alcohol-related aetiology, IRR 0.93 [95% CI 0.90–0.96] to 0.82 [95% CI 0.78–0.86]; Figures 1 and 2, Appendices 8 and 13).

Viral-related CLD accounted for 2.8–5.7% of younger and 0.5–1.1% of older cohorts (Table 1). Among younger cohorts, compared with non-viral-related CLD (uptake 30.0–39.3%), uptake was higher for viral-related aetiology (29.9–38.6%, IRR 1.38 [95% CI 1.30–1.46] to 1.64 [95% CI 1.54–1.74]) and uncertain viral aetiology (41.2–51.4%, IRR 1.28 [95% CI 1.25–1.32] to 1.47 [95% CI 1.40–1.55]; Figures 1 and 2, Appendices 7 and 14). Among older cohorts, this association attenuated and reversed, such that uptake was lowest among those with viral aetiology (55.4–72.3%) compared to non-viral-related aetiology (71.2–80.2%; IRR 0.84 [95% CI 0.74–0.95] to 1.05 [95% CI 0.97–1.13]) and uncertain viral aetiology (72.1–77.9%, IRR 0.92 [95% CI 0.89–0.94] to 0.98 [95% CI 0.96–1.00]; Figures 1 and 2, Appendices 8 and 14).

Among younger cohorts, uptake was higher for those with Green Book-listed diagnoses (35.4–48.1% vs 30.3–39.5%; IRR 1.26 [95% CI 1.22–1.29] to 1.45 [95% CI 1.39–1.51]; Figures 1 and 2, Appendices 7 and 15). In older cohorts, there was a modest reduction in uptake among individuals with listed versus non-listed diagnoses (68.8–76.2% vs 71.5–80.3%; IRR 0.96 [95% CI 0.94–0.98] to 0.88 [95% CI 0.86–0.91]; Figures 1 and 2, Appendices 8 and 15).

## Discussion

Among adults with CLD in England, uptake of seasonal influenza vaccine was 30.9–40.5% in those below the age eligibility threshold, and 71.1–79.7% in those additionally eligible due to age, highlighting a large difference in uptake between age- and clinical risk-based eligibility.

In younger individuals with CLD and no other qualifying clinical risk comorbidity, severity of CLD was an important independent determinant of vaccine uptake. In contrast, in those with at least one other clinical risk comorbidity, the effect of CLD severity on uptake was negligible. Alcohol-related and viral aetiology were also independent determinants of influenza vaccine uptake in younger cohorts. Conversely, in older individuals, increased severity, alcohol-related aetiology and viral aetiology were generally associated with a modest reduction in vaccine uptake. This effect was greatest in older individuals with CLD but no other clinical risk comorbidity. Overall, this suggests that having additional indications for influenza vaccination through age and clinical risk comorbidities attenuates (and in some cases reverses) the effect of CLD severity on uptake.

To our knowledge there are no existing studies which have investigated the effect of severity or aetiology of CLD on influenza vaccine uptake. Qualitative research in England with adults in clinical risk groups found that chronic disease management pathways did not consistently recommend influenza vaccination in outpatient consultations.^26^ Low perceived risk of influenza and limited awareness of the relevance of vaccination to chronic disease management were key reasons for vaccine non-uptake.^26^ Among younger individuals without additional clinical risk comorbidities, in whom uptake was particularly low, greater CLD severity may increase perceived risk and awareness of vaccination benefits, leading to higher uptake. This could be through more frequent healthcare contacts, which was suggested as an important determinant of influenza vaccine uptake in a Spanish study of individuals with diabetes.^27^ Individuals with more severe disease may also be more likely to be reviewed by specialists outside of primary care, which may provide additional reminders or opportunities to receive vaccination.

Younger individuals with other comorbidities had higher uptake than those with CLD alone. The attenuated effect of severity in these individuals may reflect altered perception of risk by individuals and vaccination providers, alongside increased opportunities for vaccination irrespective of CLD severity during the management of multi-morbidity. The inverse association between severity and vaccine uptake among older individuals may reflect greater barriers to accessing vaccination services by those with more complex health needs, or competing medical priorities for long-term condition management during consultations.

We updated existing codelists to define CLD and other clinical risk groups, aiming to align with Green Book criteria. However, it is possible that some of the observed associations reflect the extent to which different subcategories of CLD match the population invited for vaccination by GPs. Among younger adults, increased uptake in association with severe CLD may reflect closer alignment of more severe versus low severity categories with those invited for vaccination. Low severity mostly includes individuals with steatosis, which is not specifically listed in the Green Book. When additional clinical risk comorbidities or age-based eligibility criteria are met, individuals included in this study may be more likely to accurately reflect those invited for vaccination.

The NHS provides codelists and search criteria to identify and invite vaccine-eligible individuals enrolled at GP practices annually.^28^ Since 2022, vaccine invitations have also been sent via a centralised NHS service,^29^ although this uses distinct codelists so individuals in clinical risk groups may not align with those in GP-based searches. In addition, decisions over vaccine eligibility rely on clinical judgement. The Green Book provides examples rather than exhaustive lists of diagnoses within clinical risk groups including CLD. Furthermore, communication campaigns by the NHS and UKHSA, among others, may influence the propensity of individuals to request vaccination based on personal understanding of their clinical history. The inherent ambiguity in defining clinical risk groups presented a challenge to this study. In contrast to age-based eligibility, it also presents a challenge to the programme itself in terms of identifying individuals and communicating eligibility.

In terms of strengths, this study used primary care electronic health records which are the most complete source of exposure and influenza vaccination data available. The data extract included 24.3% of the English population and covered five consecutive influenza seasons. A low proportion of individuals exited follow-up prior to vaccination, and the only variable with missing data was ethnicity, which did not introduce selection bias. Stratified analyses helped identify different effects in important subgroups.

As for limitations, information bias may have arisen from severity classification because of the variability of severity within each individual diagnosis rather than across all diagnoses. Severity classifications exist for cirrhosis (Child-Pugh and Model for End-stage Liver Disease) but require laboratory results which were not available. Misclassification is also likely from coding inaccuracies, with more severe disease potentially recorded more reliably due to increased healthcare contact. Exposure misclassification may occur given that we did not account for potential resolution of diagnoses, which is more likely to affect milder disease. Vaccination under-ascertainment may be differential if individuals with milder disease are more often vaccinated in pharmacies. While pharmacies are expected to inform GPs of administered vaccinations,^4^ there is no up-to-date evidence quantifying how frequently this occurs.

A large proportion of individuals with CLD – particularly those under 65 years of age – are not benefiting from seasonal influenza vaccination. Providing information on the importance of influenza vaccination as part of chronic disease management should be routinely incorporated into CLD care pathways in both primary and secondary services (in outpatient settings or following acute admissions). Future research should prioritise understanding the facilitators and barriers to influenza vaccine delivery to and uptake by individuals with CLD. This should focus on younger individuals (given lower uptake) and those with more severe disease (given higher risk of morbidity). Greater clarity on vaccine eligibility could be achieved by naming additional CLD diagnoses in the Green Book, such as steatosis. As vaccine delivery increasingly occurs within pharmacy settings,^30^ research is needed to develop linked datasets to quantify the extent to which vaccinations are subsequently captured in GP records. The development of automated data transfer mechanisms, for example shared care records, would facilitate patient management and improve the accuracy of programme evaluation.

## Data Availability

This study uses data from the Clinical Practice Research Datalink (CPRD) which does not allow sharing of patient-level data. The specification for the CPRD data set used in this study is available at: https://www.cprd.com/doi/cprd-aurum-june-2024-dataset. Analysis code and code lists are shared via a Github repository available at: https://github.com/Eyedeet/Flu_uptake_England/.

## Ethical approval

Ethical approval was granted by the London School of Hygiene and Tropical Medicine’s Ethics Committee (reference 32001). Data governance approval was obtained from CPRD’s Independent Scientific Advisory Committee (protocol 22_001750).

## Contributors

Conceptualisation: ILH, SLRM, SM-J, HIM, EPKP, AMS; Methodology: ILH, DE, EPKP, AMS; Formal analysis: ILH; Writing–original draft: ILH; Writing–review & editing: ILH, DE, CNJC, SLRM, JLB, SM-J, BK-D, HIM, EPKP, AMS; Visualisation: ILH; Project administration: EPKP, AMS.

## Data sharing

This study uses data from the Clinical Practice Research Datalink (CPRD) which does not allow sharing of patient-level data. The specification for the CPRD data set used in this study is available at: https://www.cprd.com/doi/cprd-aurum-june-2024-dataset. Analysis code and code lists are shared via a Github repository available at: https://github.com/Eyedeet/Influenza_uptake_England/.

## Acknowledgements

This study is based on data from the CPRD obtained under licence from the UK Medicines & Healthcare products Regulatory Agency. The data is provided by patients and collected by the NHS as part of their care and support. We are grateful to the millions of individuals who contribute their data for the purpose of improving clinical and public health services in the UK.

This study is funded by the National Institute for Health and Care Research (NIHR) Health Protection Research Unit in Vaccines and Immunisation (grant numbers NIHR200929 and NIHR207408), a partnership between UK Health Security Agency and the London School of Hygiene and Tropical Medicine. The views expressed are those of the authors and not necessarily those of the NIHR, UK Health Security Agency, or the Department of Health and Social Care.

