## Supplementary Appendix for "The association between severity and aetiology of chronic liver disease and seasonal influenza vaccination uptake in adults: a retrospective cohort study using English primary care data (2019–2024)"

**Supplementary Material**

**Table of Contents**

Appendix 1: Data processing steps undertaken to produce the analysis dataset. 2

Appendix 2: Record statement. 6

Appendix 3: Assembly of cohorts. 10

Appendix 4: Number and proportion with additional clinical risk comorbidities, by eligibility for influenza vaccination. 11

Appendix 5: Demographic characteristics of participants in each cohort, by severity of chronic liver disease, amongst individuals who were not additionally age eligible for influenza vaccination. 12

Appendix 6: Demographic characteristics of participants in each cohort, by severity of chronic liver disease, amongst individuals who were additionally age eligible for influenza vaccination. 15

Appendix 7: Number and proportion of individuals who were influenza vaccinated in each season, amongst individuals who were not additionally age eligible for influenza vaccination. 18

Appendix 8: Number and proportion of individuals who were influenza vaccina4]ted in each season, amongst individuals who were additionally age eligible for influenza vaccination. 21

Appendix 9: Incidence rate ratios of influenza vaccine uptake by severity of chronic liver disease and age eligibility for influenza vaccination. 24

Appendix 10: Number and proportion vaccinated and incidence rate ratios of influenza vaccine uptake by severity of chronic liver disease, stratified by clinical risk 25

Appendix 11: Incidence rate ratios of influenza vaccine uptake by severity of chronic liver disease, stratified by ethnicity and age eligibility for influenza vaccination (2023/24 cohort). 26

Appendix 12: Incidence rate ratios of influenza vaccine uptake by severity of chronic liver disease, stratified by Index of Multiple Deprivation (IMD) and age eligibility for influenza vaccination (2023/24 cohort). 27

Appendix 13: Incidence rate ratios of influenza vaccine uptake by alcohol aetiology of chronic liver disease and age eligibility for influenza vaccination. 28

Appendix 14: Incidence rate ratios of influenza vaccine uptake by viral aetiology of chronic liver disease and age eligibility for influenza vaccination. 29

Appendix 15: Incidence rate ratios of influenza vaccine uptake by Green Book diagnosis and age eligibility for influenza vaccination. 30

**Appendix 1: Data processing steps undertaken to produce the analysis dataset.**

1. Defining codelists for chronic liver disease, other clinical risk comorbidities, and influenza vaccine administration (which included terms for influenza antigen, influenza infection, brand names for influenza vaccines and general vaccination administration codes). Codelists are available at https://github.com/Eyedeet/Influenza_uptake_England.

2. Identification of each season's cohort using the extract provided by CPRD Aurum. This included a series of sequential steps based on the inclusion and exclusion criteria:
   - Removing individuals who were aged <18 years before the end of study follow-up (31st March 2024) or >115 years before the start of study follow-up (1st September 2018)
   - Removing individuals who died before the start of study follow-up
   - Removing individuals whose registration ended before the start of study follow-up
   - Removing individuals who were registered for less than 1 year before the start of the final influenza season (1st September 2023)
   - Removing individuals registered at practices whose last data collection was before the start of study follow-up
   - Removing individuals who were ineligible for data linkage to Hospital Episode Statistics and Office for National Statistics mortality
   - Then, for each influenza season:
     - Removing individuals aged <18 years or >115 years at the start of the season
     - Removing individuals who were registered for less than 1 year before the start of the season
     - Removing individuals who died before the start of the season
     - Removing individuals whose registration ended before the start of the season
     - Removing individuals whose practice's last data collection date was before the start of the influenza season
     - Removing individuals who did not qualify for seasonal influenza vaccination due to absence of a recorded clinical risk diagnosis or not fulfilling age eligibility criteria (≥65 years in 2019/20, 2020/21, 2023/24, or ≥50 years in 2021/22 and 2022/23)
     - Removing individuals who were registered at a duplicate practice according to recommendations by CPRD[1]

3. Processing demographic and clinical variables according to the criteria below.

| **Demographic variable** | **Values** | **Look-back period** | **Notes** |
|---|---|---|---|
| Age | Age as integer then grouped as: <40, 40–49, 50–64, 65–69, and ≥70 years | Age at start of each cohort's follow-up period (1st September) | Month and year of birth supplied by CPRD. Day of birth was assumed as 1st July for all individuals. |
| Sex | Male, female, indeterminate | – | As recorded in medical record. The currently recorded sex was assumed to apply to each individual in all five cohorts. |
| Ethnicity | White, Black, Asian, Mixed, Other | – | The most common ethnicity (excluding 'not stated') was assigned; if there were equally common ethnicities, the most recent ethnicity was assigned. If there was no ethnicity recorded, assigned 'not stated'. |

| | | | |
|---|---|---|---|
| Geographical region | London, North West, South East, West Midlands, South West, East of England, North East, Yorkshire & The Humber | – | Based on primary care practice postcode. The geographical region was assumed to apply in all five cohorts (based on most recent primary care practice postcode). |
| Index of Multiple Deprivation (IMD) | 1 (most deprived), 2, 3, 4, 5 (least deprived) | – | From small area data (linked to primary care data). Based on individual's registered postcode. If individual linkage could not be performed, IMD based on practice postcode was used. Small area data IMD values were produced in 2019 by the Ministry of Housing, Communities and Local Government (most recent version).[2] The IMD was assumed to apply in all five cohorts (based on most recent household postcode). |
| **Clinical comorbidities** | **Values** | **Look-back period** | **Notes** |
| Chronic heart disease | Yes, no | Ever recorded prior to each season's cohort | – |
| Chronic respiratory disease | Yes, no | Ever recorded prior to each season's cohort | – |
| Chronic neurological disease | Yes, no | Ever recorded prior to each season's cohort | – |
| Asthma | Yes, no | 5 years prior to each season's cohort | – |
| Chronic kidney disease | Yes, no | Ever recorded prior to each season's cohort | – |
| Diabetes | Yes, no | Ever recorded prior to each season's cohort | – |
| Immunosuppression | Yes, no | Targeted immunotherapy (i.e., biologics or small molecule drugs) or non-biological immune modulating drugs: 6 months prior to each season's cohort<br><br>Oncological diagnoses: 1 year prior to each season's cohort | – |

| | | | |
|---|---|---|---|
| | | Haematological diagnoses: ever recorded prior to each season's cohort<br><br>Other immunosuppressive diagnoses: ever recorded prior to each season's cohort | |
| Asplenia | Yes, no | Ever recorded prior to each season's cohort | – |
| Severe obesity | Yes, no | Most recent anthropometric measurements from 5 years prior to each season's cohort | Body mass index of ≥40kg/m$^2$ defined severe obesity. Measurements excluded if height <100cm or >250cm, weight <30kg or >300kg, BMI <10kg/m$^2$ or >60kg/m$^2$. BMI was calculated using height and weight measurements if a BMI value was not available. The most recent BMI measurement (or calculation) was used. |

| **Exposure** | **Values** | **Look-back period** | **Notes** |
|---|---|---|---|
| Chronic liver disease: severity | Low, moderate, severe | Ever recorded prior to each season's cohort | If more than one relevant code, the most severe classification during each cohort was used. |
| Chronic liver disease: alcohol-related aetiology | Not alcohol-related, uncertain if alcohol-related, alcohol-related | Ever recorded prior to each season's cohort | Diagnoses which included alcohol in the clinical code were classified as alcohol-related. Diagnoses which may or may not be alcohol-related were classified as uncertain.<br><br>If there was more than one relevant code which resulted in inconsistent classification between alcohol-related or uncertain/not alcohol-related, exposure classified as alcohol-related. |
| Chronic liver disease: viral-related aetiology | Not viral-related, uncertain if viral-related, viral-related | Ever recorded prior to each season's cohort | Diagnoses which included viral in the clinical code were classified as viral-related. Diagnoses which may or may not be viral-related were classified as uncertain.<br><br>If there was more than one relevant code which resulted in inconsistent classification between viral-related or uncertain/not viral-related, exposure classified as viral-related. |
| Chronic liver disease: Green Book listed diagnosis | Yes, no | Ever recorded prior to each season's cohort | Three specific diagnoses listed: cirrhosis, biliary atresia, chronic hepatitis. |

| Outcome | Values | Look-back period | Notes |
|---|---|---|---|
| Influenza vaccination | Vaccinated, unvaccinated | For each season's cohort between 1st September–28th February | Prescription and observation data tables used. Codelists incorporating terms for influenza antigen, influenza infection, and vaccine brand names. Validated algorithm applied for codes implying 'vaccine given/administered', neutral codes, vaccine product codes, and vaccine declined codes.[3]<br><br>All influenza vaccine formulations licensed for adults were included. If there was no record of an influenza vaccine being administered within a season, it was assumed not given. Where there was more than one record, date of administration was taken as the first recorded date. |

### Appendix 2: Record statement.

| | Item No. | STROBE items | Location in manuscript where items are reported | RECORD items | Location in manuscript where items are reported |
|---|---|---|---|---|---|
| **Title and abstract** | | | | | |
| | 1 | (a) Indicate the study's design with a commonly used term in the title or the abstract (b) Provide in the abstract an informative and balanced summary of what was done and what was found | Title, Abstract | RECORD 1.1: The type of data used should be specified in the title or abstract. When possible, the name of the databases used should be included.<br><br>RECORD 1.2: If applicable, the geographic region and timeframe within which the study took place should be reported in the title or abstract.<br><br>RECORD 1.3: If linkage between databases was conducted for the study, this should be clearly stated in the title or abstract. | Title, Abstract<br><br>Title, Abstract<br><br>Methods (established linkage to death data and secondary care data) |
| **Introduction** | | | | | |
| Background rationale | 2 | Explain the scientific background and rationale for the investigation being reported | Background | | |
| Objectives | 3 | State specific objectives, including any prespecified hypotheses | Background | | |
| **Methods** | | | | | |
| Study Design | 4 | Present key elements of study design early in the paper | Abstract, Methods (Study design and population) | | |
| Setting | 5 | Describe the setting, locations, and relevant dates, including periods of recruitment, exposure, follow-up, and data collection | Methods (Data sources, Study design and population, Exposures) | | |
| Participants | 6 | *(a) Cohort study* - Give the eligibility criteria, and the sources and methods of selection of participants. Describe methods of follow-up *Case-control study* - Give the eligibility criteria, and the sources and methods of case ascertainment and control selection. Give the rationale for the choice of cases and controls *Cross-sectional study* - Give the eligibility criteria, and the sources and methods of selection of participants<br><br>*(b) Cohort study* - For matched studies, give matching criteria and number of exposed and unexposed *Case-control study* - For matched studies, give matching criteria and the number of controls per case | Methods (Data sources, Study design and population, Statistical analysis)<br><br>N/A | RECORD 6.1: The methods of study population selection (such as codes or algorithms used to identify subjects) should be listed in detail. If this is not possible, an explanation should be provided.<br><br>RECORD 6.2: Any validation studies of the codes or algorithms used to select the population should be referenced. If validation was conducted for this study and not published elsewhere, detailed methods and results should be provided.<br><br>RECORD 6.3: If the study involved linkage of databases, consider use of a flow diagram or other graphical display to demonstrate the data linkage process, including the number of individuals with linked data at each stage. | Methods, Appendix 1, Github repository<br><br>Referenced: Suffel et al, 2024. DOI 0.1002/pds.5848<br><br>Linkage to secondary care and death data escribed in Methods (established linkages) |

| | | | | | |
|---|---|---|---|---|---|
| Variables | 7 | Clearly define all outcomes, exposures, predictors, potential confounders, and effect modifiers. Give diagnostic criteria, if applicable. | Exposure, Outcome, Covariates | RECORD 7.1: A complete list of codes and algorithms used to classify exposures, outcomes, confounders, and effect modifiers should be provided. If these cannot be reported, an explanation should be provided. | Exposure, Outcome, Covariates, Appendix 1, Github repository |
| Data sources/ measurement | 8 | For each variable of interest, give sources of data and details of methods of assessment (measurement).<br>Describe comparability of assessment methods if there is more than one group | Exposure, Outcome, Covariates, Appendix 1<br><br>N/A | | |
| Bias | 9 | Describe any efforts to address potential sources of bias | Statistical analysis (sequential modelling approach and sensitivity analysis) | | |
| Study size | 10 | Explain how the study size was arrived at | Appendix 3 | | |
| Quantitative variables | 11 | Explain how quantitative variables were handled in the analyses. If applicable, describe which groupings were chosen, and why | Exposure, Outcome, Covariates, Appendix 1 | | |
| Statistical methods | 12 | (a) Describe all statistical methods, including those used to control for confounding<br>(b) Describe any methods used to examine subgroups and interactions<br>(c) Explain how missing data were addressed<br>(d) *Cohort study* - If applicable, explain how loss to follow-up was addressed<br>*Case-control study* - If applicable, explain how matching of cases and controls was addressed<br>*Cross-sectional study* - If applicable, describe analytical methods taking account of sampling strategy<br>(e) Describe any sensitivity analyses | Statistical analysis | | |
| Data access and cleaning methods | | .. | | RECORD 12.1: Authors should describe the extent to which the investigators had access to the database population used to create the study population.<br><br>RECORD 12.2: Authors should provide information on the data cleaning methods used in the study. | Appendix 3, Data sharing<br><br>Appendix 3 |
| Linkage | | .. | | RECORD 12.3: State whether the study included person-level, institutional-level, or other data linkage across two or more databases. The methods of linkage and methods of linkage quality evaluation should be provided. | Data sources |
| **Results** | | | | | |
| Participants | 13 | (a) Report the numbers of individuals at each stage of the study (*e.g.*, numbers potentially eligible, examined for eligibility, confirmed | Appendix 3 | RECORD 13.1: Describe in detail the selection of the persons included in the study (*i.e.,* study population selection) including filtering based on data quality, data availability and linkage. The selection of included | Appendix 2 |

| | | | | | |
|---|---|---|---|---|---|
| | | eligible, included in the study, completing follow-up, and analysed)<br>(b) Give reasons for non-participation at each stage.<br>(c) Consider use of a flow diagram | | persons can be described in the text and/or by means of the study flow diagram. | |
| Descriptive data | 14 | (a) Give characteristics of study participants (*e.g.*, demographic, clinical, social) and information on exposures and potential confounders<br>(b) Indicate the number of participants with missing data for each variable of interest<br>(c) *Cohort study* - summarise follow-up time (*e.g.*, average and total amount) | Results, Table 1, Appendix 4, Appendix 5, 6 | | |
| Outcome data | 15 | *Cohort study* - Report numbers of outcome events or summary measures over time<br>*Case-control study* - Report numbers in each exposure category, or summary measures of exposure<br>*Cross-sectional study* - Report numbers of outcome events or summary measures | Appendix 7, Appendix 8 | | |
| Main results | 16 | (a) Give unadjusted estimates and, if applicable, confounder-adjusted estimates and their precision (e.g., 95% confidence interval). Make clear which confounders were adjusted for and why they were included<br>(b) Report category boundaries when continuous variables were categorized<br>(c) If relevant, consider translating estimates of relative risk into absolute risk for a meaningful time period | Figure 2, Appendix 9, Appendix 13, Appendix 14, Appendix 15 | | |
| Other analyses | 17 | Report other analyses done—e.g., analyses of subgroups and interactions, and sensitivity analyses | Appendix 10, Appendix 11, Appendix 12, | | |
| **Discussion** | | | | | |
| Key results | 18 | Summarise key results with reference to study objectives | Discussion (first paragraph) | | |
| Limitations | 19 | Discuss limitations of the study, taking into account sources of potential bias or imprecision. Discuss both direction and magnitude of any potential bias | Discussion (limitations paragraph) | RECORD 19.1: Discuss the implications of using data that were not created or collected to answer the specific research question(s). Include discussion of misclassification bias, unmeasured confounding, missing data, and changing eligibility over time, as they pertain to the study being reported. | Discussion (limitations paragraph) |
| Interpretation | 20 | Give a cautious overall interpretation of results considering objectives, limitations, multiplicity of analyses, results from similar studies, and other relevant evidence | Discussion | | |
| Generalisability | 21 | Discuss the generalisability (external validity) of the study results | Not discussed | | |

| Other Information | | | | | |
|---|---|---|---|---|---|
| Funding | 22 | Give the source of funding and the role of the funders for the present study and, if applicable, for the original study on which the present article is based | Abstract, Acknowledgements, Funding | | |
| Accessibility of protocol, raw data, and programming code | | .. | | RECORD 22.1: Authors should provide information on how to access any supplemental information such as the study protocol, raw data, or programming code. | Link provided for Github repository containing codelists and code |

The REporting of studies Conducted using Observational Routinely-collected health Data (RECORD) Statement.[4]

**Appendix 3: Assembly of cohorts.**

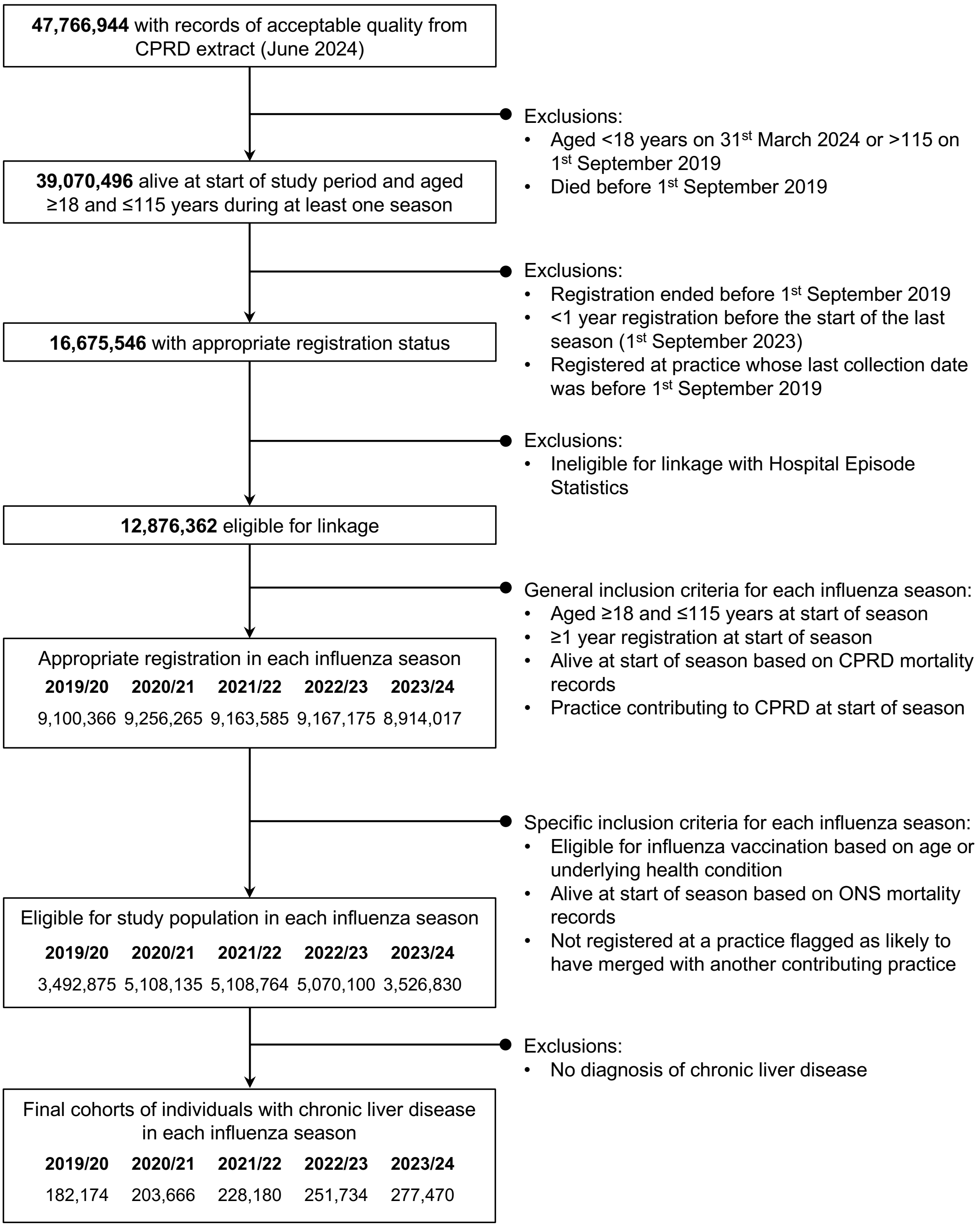

**Appendix 4: Number and proportion with additional clinical risk comorbidities, by eligibility for influenza vaccination.**

| | Eligible due to chronic liver disease (not additionally age eligible) | | | | | Eligible due to chronic liver disease and age | | | | |
|---|---|---|---|---|---|---|---|---|---|---|
| **Additional clinical risk comorbidity** | **2019/20** 122,161 (100.0)[1] | **2020/21** 56,890 (100.0)[1] | **2021/22** 63,156 (100.0)[1] | **2022/23** 69,789 (100.0)[1] | **2023/24** 183,083 (100.0)[1] | **2019/20** 60,013 (100.0)[1] | **2020/21** 146,776 (100.0)[1] | **2021/22** 165,024 (100.0)[1] | **2022/23** 181,945 (100.0)[1] | **2023/2024** 94,387 (100.0)[1] |
| **Diabetes** | 28,862 (23.6) | 8,612 (15.1) | 9,874 (15.6) | 10,874 (15.6) | 44,885 (24.5) | 23,765 (39.6) | 50,105 (34.1) | 57,090 (34.6) | 63,397 (34.8) | 37,897 (40.2) |
| **Asthma** | 17,327 (14.2) | 7,428 (13.1) | 8,275 (13.1) | 9,209 (13.2) | 26,361 (14.4) | 8,682 (14.5) | 21,808 (14.9) | 24,597 (14.9) | 27,110 (14.9) | 14,057 (14.9) |
| **Immunosuppression** | 8,411 (6.9) | 2,886 (5.1) | 3,082 (4.9) | 3,344 (4.8) | 11,706 (6.4) | 7,063 (11.8) | 13,808 (9.4) | 15,546 (9.4) | 17,185 (9.4) | 10,670 (11.3) |
| **Obesity[2]** | 9,757 (8.0) | 5,005 (8.8) | 5,797 (9.2) | 6,655 (9.5) | 16,785 (9.2) | 2,827 (4.7) | 9,608 (6.5) | 11,253 (6.8) | 12,647 (7.0) | 5,067 (5.4) |
| **Cardiovascular disease** | 7,643 (6.3) | 1,240 (2.2) | 1,424 (2.3) | 1,570 (2.2) | 11,807 (6.4) | 14,075 (23.5) | 23,183 (15.8) | 26,043 (15.8) | 28,976 (15.9) | 22,203 (23.5) |
| **Respiratory disease** | 6,649 (5.4) | 1,162 (2.0) | 1,231 (1.9) | 1,311 (1.9) | 9,238 (5.0) | 9,476 (15.8) | 16,609 (11.3) | 18,312 (11.1) | 20,107 (11.1) | 14,510 (15.4) |
| **Neurological disease** | 4,416 (3.6) | 1,831 (3.2) | 2,161 (3.4) | 2,443 (3.5) | 7,221 (3.9) | 5,129 (8.5) | 8,874 (6.0) | 10,058 (6.1) | 11,472 (6.3) | 8,464 (9.0) |
| **Renal disease** | 3,432 (2.8) | 503 (0.9) | 509 (0.8) | 562 (0.8) | 4,974 (2.7) | 11,798 (19.7) | 16,006 (10.9) | 18,107 (11.0) | 20,309 (11.2) | 18,658 (19.8) |

[1]n (column %)

[2]Body mass index ≥40kg/m$^2$

In 2019/20 and 2023/24 individuals aged ≥65 years were eligible for influenza vaccination (irrespective of additional clinical risk comorbidities). In 2020/21, 2021/22, and 2022/23 age eligibility was widened to ≥50 years due to the COVID-19 pandemic.

**Appendix 5: Demographic characteristics of participants in each cohort, by severity of chronic liver disease, amongst individuals who were not additionally age eligible for influenza vaccination.**

| | 2019/20 | | | 2020/21 | | | 2021/22 | | | 2022/23 | | | 2023/24 | | |
|---|---|---|---|---|---|---|---|---|---|---|---|---|---|---|---|
| | **Low** 98,453 (80.6)[1] | **Moderate** 11,170 (9.1)[1] | **Severe** 12,538 (10.3)[1] | **Low** 46,413 (81.6)[1] | **Moderate** 6,854 (12.0)[1] | **Severe** 3,623 (6.4)[1] | **Low** 52,625 (83.3)[1] | **Moderate** 6,914 (10.9)[1] | **Severe** 3,617 (5.7)[1] | **Low** 59,137 (84.7)[1] | **Moderate** 7,019 (10.1)[1] | **Severe** 3,633 (5.21)[1] | **Low** 157,058 (85.8)[1] | **Moderate** 12,328 (6.7)[1] | **Severe** 13,697 (7.5)[1] |
| **Age group (years)** | | | | | | | | | | | | | | | |
| <40 | 16,790 (78.9) | 3,320 (15.6) | 1,167 (5.5) | 19,155 (80.4) | 3,474 (14.6) | 1,192 (5.0) | 21,992 (82.8) | 3,399 (12.8) | 1,182 (4.4) | 25,121 (84.6) | 3,376 (11.4) | 1,210 (4.1) | 27,661 (86.4) | 3,161 (9.9) | 1,210 (3.8) |
| 40-49 | 24,357 (81.4) | 3,142 (10.5) | 2,428 (8.1) | 27,258 (82.4) | 3,380 (10.2) | 2,431 (7.4) | 30,633 (83.7) | 3,515 (9.6) | 2,435 (6.7) | 34,016 (84.9) | 3,643 (9.1) | 2,423 (6.0) | 37,828 (86.1) | 3,617 (8.2) | 2,491 (5.7) |
| 50-64 | 57,306 (80.8) | 4,708 (6.6) | 8,943 (12.6) | – | – | – | – | – | – | – | – | – | 91,569 (85.5) | 5,550 (5.2) | 9,996 (9.3) |
| 65-69 | – | – | – | – | – | – | – | – | – | – | – | – | – | – | – |
| ≥70 | – | – | – | – | – | – | – | – | – | – | – | – | – | – | – |
| **Sex** | | | | | | | | | | | | | | | |
| Male | 55,667 (80.4) | 6,320 (9.1) | 7,209 (10.4) | 28,674 (83.1) | 3,778 (10.9) | 2,056 (6.0) | 31,808 (84.5) | 3,766 (10.0) | 2,054 (5.5) | 35,346 (85.8) | 3,765 (9.1) | 2,080 (5.0) | 85,439 (85.3) | 6,903 (6.9) | 7,823 (7.8) |
| Female[2] | – (80.8) | – (9.2) | – (10.1) | – (79.3) | – (13.7) | – (7.0) | – (81.5) | – (12.3) | – (6.1) | – (83.2) | – (11.4) | – (5.4) | – (86.4) | – (6.5) | – (7.1) |
| Indetermined[3] | <10 | <10 | <10 | <10 | <10 | <10 | <10 | <10 | <10 | <10 | <10 | <10 | <10 | <10 | <10 |
| **Ethnicity** | | | | | | | | | | | | | | | |
| White | 63,294 (81.7) | 5,411 (7.0) | 8,806 (11.4) | 27,009 (83.6) | 2,884 (8.9) | 2,432 (7.5) | 31,984 (85.1) | 3,061 (8.1) | 2,558 (6.8) | 37,689 (86.5) | 3,168 (7.3) | 2,695 (6.2) | 113,242 (86.5) | 6,583 (5.0) | 11,086 (8.5) |
| Asian | 13,921 (87.5) | 1,227 (7.7) | 756 (4.8) | 9,137 (88.6) | 894 (8.7) | 287 (2.8) | 10,366 (89.6) | 903 (7.8) | 306 (2.6) | 11,803 (90.0) | 961 (7.3) | 349 (2.7) | 23,476 (90.8) | 1,438 (5.6) | 949 (3.7) |
| Black | 3,820 (64.0) | 1,741 (29.2) | 404 (6.8) | 1,759 (55.3) | 1,252 (39.4) | 167 (5.3) | 2,032 (58.1) | 1,287 (36.8) | 180 (5.1) | 2,383 (61.1) | 1,335 (34.2) | 181 (4.6) | 6,770 (71.8) | 2,155 (22.9) | 505 (5.4) |

| | 2019/20 | | | 2020/21 | | | 2021/22 | | | 2022/23 | | | 2023/24 | | |
|---|---|---|---|---|---|---|---|---|---|---|---|---|---|---|---|
| | **Low** 98,453 (80.6)[1] | **Moderate** 11,170 (9.1)[1] | **Severe** 12,538 (10.3)[1] | **Low** 46,413 (81.6)[1] | **Moderate** 6,854 (12.0)[1] | **Severe** 3,623 (6.4)[1] | **Low** 52,625 (83.3)[1] | **Moderate** 6,914 (10.9)[1] | **Severe** 3,617 (5.7)[1] | **Low** 59,137 (84.7)[1] | **Moderate** 7,019 (10.1)[1] | **Severe** 3,633 (5.21)[1] | **Low** 157,058 (85.8)[1] | **Moderate** 12,328 (6.7)[1] | **Severe** 13,697 (7.5)[1] |
| Other | 2,499 (70.2) | 894 (25.1) | 167 (4.7) | 1,567 (67.3) | 704 (30.2) | 59 (2.5) | 1,768 (69.3) | 720 (28.2) | 64 (2.5) | 2,120 (71.6) | 774 (26.1) | 66 (2.2) | 4,683 (77.3) | 1,131 (18.7) | 246 (4.1) |
| Mixed | 1,397 (75.5) | 337 (18.2) | 117 (6.3) | 789 (73.3) | 239 (22.2) | 48 (4.5) | 926 (76.6) | 238 (19.7) | 45 (3.7) | 1,082 (78.5) | 248 (18.0) | 48 (3.5) | 2,491 (82.1) | 389 (12.8) | 153 (5.0) |
| Missing | 13,522 (77.8) | 1,560 (9.0) | 2,288 (13.2) | 6,152 (80.3) | 881 (11.5) | 630 (8.2) | 5,549 (82.6) | 705 (10.5) | 464 (6.9) | 4,060 (83.1) | 533 (10.9) | 294 (6.0) | 6,396 (82.1) | 632 (8.1) | 758 (9.7) |
| **Region** | | | | | | | | | | | | | | | |
| London | 25,805 (80.6) | 3,877 (12.1) | 2,335 (7.3) | 13,261 (80.6) | 2,456 (14.9) | 739 (4.5) | 14,672 (82.2) | 2,437 (13.7) | 732 (4.1) | 16,517 (83.6) | 2,499 (12.6) | 739 (3.7) | 40,095 (85.6) | 4,201 (9.0) | 2,522 (5.4) |
| North West | 18,839 (79.6) | 2,038 (8.6) | 2,793 (11.8) | 8,895 (81.8) | 1,164 (10.7) | 815 (7.5) | 10,268 (83.8) | 1,172 (9.6) | 811 (6.6) | 11,727 (85.2) | 1,217 (8.8) | 813 (5.9) | 32,363 (85.3) | 2,316 (6.1) | 3,268 (8.6) |
| South East | 16,904 (81.4) | 1,681 (8.1) | 2,191 (10.5) | 7,942 (83.3) | 991 (10.4) | 601 (6.3) | 9,292 (85.1) | 1,035 (9.5) | 595 (5.4) | 10,678 (86.5) | 1,041 (8.4) | 624 (5.1) | 29,339 (87.4) | 1,891 (5.6) | 2,347 (7.0) |
| West Midlands | 13,755 (80.1) | 1,401 (8.2) | 2,019 (11.8) | 6,497 (81.3) | 945 (11.8) | 547 (6.8) | 7,319 (82.9) | 949 (10.8) | 557 (6.3) | 8,531 (84.3) | 987 (9.8) | 603 (6.0) | 22,708 (85.2) | 1,598 (6.0) | 2,351 (8.8) |
| South West | 10,625 (79.4) | 1,139 (8.5) | 1,619 (12.1) | 4,540 (80.3) | 660 (11.7) | 456 (8.1) | 4,988 (81.5) | 681 (11.1) | 453 (7.4) | 5,180 (82.6) | 665 (10.6) | 426 (6.8) | 14,064 (82.6) | 1,296 (7.6) | 1,668 (9.8) |
| East of England | 3,114 (82.4) | 282 (7.5) | 384 (10.2) | 1,390 (83.9) | 176 (10.6) | 91 (5.5) | 1,812 (86.8) | 172 (8.2) | 103 (4.9) | 2,204 (88.1) | 193 (7.7) | 106 (4.2) | 6,763 (90.1) | 336 (4.5) | 408 (5.4) |
| North East | 3,866 (82.5) | 250 (5.3) | 572 (12.2) | 1,559 (81.9) | 163 (8.6) | 182 (9.6) | 1,743 (83.7) | 160 (7.7) | 180 (8.6) | 1,919 (85.2) | 148 (6.6) | 186 (8.3) | 5,360 (85.3) | 287 (4.6) | 640 (10.2) |
| Yorkshire & The Humber | 3,108 (82.1) | 297 (7.8) | 379 (10.0) | 1,337 (81.2) | 179 (10.9) | 131 (8.0) | 1,514 (82.3) | 194 (10.5) | 132 (7.2) | 1,417 (85.0) | 162 (9.7) | 89 (5.3) | 3,812 (87.4) | 255 (5.8) | 295 (6.8) |
| East Midlands | 2,437 (84.4) | 205 (7.1) | 246 (8.5) | 992 (84.6) | 120 (10.2) | 61 (5.2) | 1,017 (85.8) | 114 (9.6) | 54 (4.6) | 964 (86.2) | 107 (9.6) | 47 (4.2) | 2,554 (88.1) | 148 (5.1) | 198 (6.8) |

| | 2019/20 | | | 2020/21 | | | 2021/22 | | | 2022/23 | | | 2023/24 | | |
|---|---|---|---|---|---|---|---|---|---|---|---|---|---|---|---|
| | **Low** 98,453 (80.6)[1] | **Moderate** 11,170 (9.1)[1] | **Severe** 12,538 (10.3)[1] | **Low** 46,413 (81.6)[1] | **Moderate** 6,854 (12.0)[1] | **Severe** 3,623 (6.4)[1] | **Low** 52,625 (83.3)[1] | **Moderate** 6,914 (10.9)[1] | **Severe** 3,617 (5.7)[1] | **Low** 59,137 (84.7)[1] | **Moderate** 7,019 (10.1)[1] | **Severe** 3,633 (5.21)[1] | **Low** 157,058 (85.8)[1] | **Moderate** 12,328 (6.7)[1] | **Severe** 13,697 (7.5)[1] |
| **Socioeconomic status[4]** | | | | | | | | | | | | | | | |
| 1 (Least deprived) | 15,530 (84.0) | 1,303 (7.0) | 1,666 (9.0) | 6,285 (84.4) | 716 (9.6) | 446 (6.0) | 7,168 (85.8) | 750 (9.0) | 438 (5.2) | 8,068 (87.0) | 768 (8.3) | 440 (4.7) | 24,706 (88.2) | 1,445 (5.2) | 1,858 (6.6) |
| 2 | 16,804 (83.2) | 1,450 (7.2) | 1,944 (9.6) | 7,189 (84.1) | 869 (10.2) | 492 (5.8) | 8,102 (85.3) | 900 (9.5) | 499 (5.3) | 9,082 (86.4) | 937 (8.9) | 488 (4.6) | 26,592 (87.6) | 1,696 (5.6) | 2,075 (6.8) |
| 3 | 18,659 (81.6) | 1,917 (8.4) | 2,295 (10.0) | 8,674 (82.7) | 1,177 (11.2) | 637 (6.1) | 9,703 (84.0) | 1,176 (10.2) | 666 (5.8) | 10,870 (85.3) | 1,207 (9.5) | 672 (5.3) | 29,673 (86.6) | 2,088 (6.1) | 2,487 (7.3) |
| 4 | 22,994 (79.5) | 3,020 (10.4) | 2,894 (10.0) | 11,520 (80.6) | 1,898 (13.3) | 868 (6.1) | 13,104 (82.6) | 1,887 (11.9) | 876 (5.5) | 14,727 (84.2) | 1,879 (10.7) | 875 (5.0) | 36,929 (85.0) | 3,268 (7.5) | 3,234 (7.4) |
| 5 (Most deprived) | 24,466 (77.2) | 3,480 (11.0) | 3,739 (11.8) | 12,745 (79.1) | 2,194 (13.6) | 1,180 (7.3) | 14,548 (81.3) | 2,201 (12.3) | 1,138 (6.4) | 16,390 (82.9) | 2,228 (11.3) | 1,158 (5.9) | 39,158 (83.3) | 3,831 (8.1) | 4,043 (8.6) |
| **Has another clinical risk comorbidity** | | | | | | | | | | | | | | | |
| No | 50,634 (80.3) | 7,232 (11.5) | 5,199 (8.2) | 28,384 (80.3) | 5,185 (14.7) | 1,765 (5.0) | 31,844 (81.9) | 5,236 (13.5) | 1,784 (4.6) | 35,855 (83.5) | 5,291 (12.3) | 1,772 (4.1) | 79,218 (85.5) | 7,940 (8.6) | 5,530 (6.0) |
| Yes | 47,819 (80.9) | 3,938 (6.7) | 7,339 (12.4) | 18,029 (83.6) | 1,669 (7.7) | 1,858 (8.6) | 20,781 (85.5) | 1,678 (6.9) | 1,833 (7.5) | 23,282 (86.6) | 1,728 (6.4) | 1,861 (6.9) | 77,840 (86.1) | 4,388 (4.9) | 8,167 (9.0) |

[1]n (row %)

[2]Number suppressed so that number of individuals with indetermined sex cannot be calculated.

[3]Number and proportions suppressed to avoid statistical disclosure.

[4]Measured by Index of Multiple Deprivation.

In 2019/20 and 2023/24 individuals aged ≥65 years were eligible for influenza vaccination (irrespective of additional clinical risk comorbidities). In 2020/21, 2021/22, and 2022/23 age eligibility was widened to ≥50 years due to the COVID-19 pandemic.

**Appendix 6: Demographic characteristics of participants in each cohort, by severity of chronic liver disease, amongst individuals who were additionally age eligible for influenza vaccination.**

| | 2019/20 | | | 2020/21 | | | 2021/22 | | | 2022/23 | | | 2023/24 | | |
|---|---|---|---|---|---|---|---|---|---|---|---|---|---|---|---|
| | **Low** 45,810 (76.3)[1] | **Moderate** 3,544 (5.9)[1] | **Severe** 10,659 (17.8)[1] | **Low** 117,328 (79.9)[1] | **Moderate** 8,786 (6.0)[1] | **Severe** 20,662 (14.1)[1] | **Low** 134,204 (81.3)[1] | **Moderate** 9,303 (5.6)[1] | **Severe** 21,517 (13.0)[1] | **Low** 150,090 (82.5)[1] | **Moderate** 9,671 (5.32)[1] | **Severe** 22,184 (12.2)[1] | **Low** 76,888 (81.5)[1] | **Moderate** 4,591 (4.9)[1] | **Severe** 12,908 (13.7)[1] |
| **Age group (years)** | | | | | | | | | | | | | | | |
| <40 | — | — | — | — | — | — | — | — | — | — | — | — | — | — | — |
| 40-49 | — | — | — | — | — | — | — | — | — | — | — | — | — | — | — |
| 50-64 | — | — | — | 64,990 (81.9) | 4,988 (6.3) | 9,399 (11.8) | 74,199 (83.3) | 5,199 (5.8) | 9,684 (10.9) | 82,533 (84.4) | 5,369 (5.5) | 9,884 (10.1) | — | — | — |
| 65-69 | 16,192 (78.8) | 1,073 (5.2) | 3,296 (16.0) | 18,140 (80.1) | 1,152 (5.1) | 3,366 (14.9) | 20,679 (81.3) | 1,279 (5.0) | 3,491 (13.7) | 23,047 (82.1) | 1,318 (4.7) | 3,714 (13.2) | 26,275 (83.3) | 1,439 (4.6) | 3,811 (12.1) |
| ≥70 | 29,618 (75.1) | 2,471 (6.3) | 7,363 (18.7) | 34,198 (76.4) | 2,646 (5.9) | 7,897 (17.7) | 39,326 (77.9) | 2,825 (5.6) | 8,342 (16.5) | 44,510 (79.4) | 2,984 (5.3) | 8,586 (15.3) | 50,613 (80.5) | 3,152 (5.0) | 9,097 (14.5) |
| **Sex** | | | | | | | | | | | | | | | |
| Male | 20,991 (75.6) | 1,557 (5.6) | 5,216 (18.8) | 58,078 (78.8) | 4,625 (6.3) | 10,962 (14.9) | 65,970 (80.1) | 4,895 (5.9) | 11,448 (13.9) | 73,351 (81.1) | 5,152 (5.7) | 11,903 (13.2) | 35,287 (80.3) | 2,181 (5.0) | 6,502 (14.8) |
| Female[2] | — (77.0) | — (6.2) | — (16.9) | — (81.0) | — (5.7) | — (13.3) | — (82.5) | — (5.3) | — (12.2) | — (83.8) | — (4.9) | — (11.2) | — (82.5) | — (4.8) | — (12.7) |
| Indetermined[3] | <10 | <10 | <10 | <10 | <10 | <10 | <10 | <10 | <10 | <10 | <10 | <10 | <10 | <10 | <10 |
| **Ethnicity** | | | | | | | | | | | | | | | |
| White | 34,587 (76.7) | 2,505 (5.6) | 7,995 (17.7) | 84,449 (80.2) | 5,586 (5.3) | 15,268 (14.5) | 102,148 (81.4) | 6,192 (4.9) | 17,094 (13.6) | 119,326 (82.5) | 6,689 (4.6) | 18,590 (12.9) | 65,270 (81.3) | 3,674 (4.6) | 11,322 (14.1) |
| Asian | 2,964 (84.0) | 158 (4.5) | 405 (11.5) | 10,356 (86.9) | 592 (5.0) | 966 (8.1) | 11,880 (87.7) | 671 (5.0) | 1,001 (7.4) | 13,984 (88.2) | 747 (4.7) | 1,126 (7.1) | 5,730 (86.4) | 296 (4.5) | 605 (9.1) |
| Black | 1,039 (75.1) | 150 (10.8) | 194 (14.0) | 3,817 (75.2) | 802 (15.8) | 457 (9.0) | 4,491 (76.3) | 905 (15.4) | 488 (8.3) | 5,264 (77.6) | 1,000 (14.7) | 516 (7.6) | 2,043 (81.2) | 235 (9.3) | 238 (9.5) |

| | 2019/20 | | | 2020/21 | | | 2021/22 | | | 2022/23 | | | 2023/24 | | |
|---|---|---|---|---|---|---|---|---|---|---|---|---|---|---|---|
| | **Low** 45,810 (76.3)[1] | **Moderate** 3,544 (5.9)[1] | **Severe** 10,659 (17.8)[1] | **Low** 117,328 (79.9)[1] | **Moderate** 8,786 (6.0)[1] | **Severe** 20,662 (14.1)[1] | **Low** 134,204 (81.3)[1] | **Moderate** 9,303 (5.6)[1] | **Severe** 21,517 (13.0)[1] | **Low** 150,090 (82.5)[1] | **Moderate** 9,671 (5.32)[1] | **Severe** 22,184 (12.2)[1] | **Low** 76,888 (81.5)[1] | **Moderate** 4,591 (4.9)[1] | **Severe** 12,908 (13.7)[1] |
| Other | 467 (72.3) | 82 (12.7) | 97 (15.0) | 1,979 (77.3) | 354 (13.8) | 228 (8.9) | 2,368 (77.8) | 418 (13.7) | 256 (8.4) | 2,889 (79.5) | 468 (12.9) | 277 (7.6) | 1,167 (77.9) | 178 (11.9) | 153 (10.2) |
| Mixed | 235 (73.7) | 31 (9.7) | 53 (16.6) | 1,075 (78.5) | 157 (11.5) | 137 (10.0) | 1,265 (80.1) | 176 (11.1) | 138 (8.7) | 1,547 (81.3) | 191 (10.0) | 164 (8.6) | 556 (80.6) | 59 (8.6) | 75 (10.9) |
| Missing | 6,518 (72.0) | 618 (6.8) | 1,915 (21.2) | 15,652 (76.2) | 1,295 (6.3) | 3,606 (17.5) | 12,052 (77.6) | 941 (6.1) | 2,540 (16.4) | 7,080 (77.2) | 576 (6.3) | 1,511 (16.5) | 2,122 (76.2) | 149 (5.3) | 515 (18.5) |
| **Region** | | | | | | | | | | | | | | | |
| London | 8,961 (79.6) | 695 (6.2) | 1,605 (14.3) | 25,704 (81.8) | 2,349 (7.5) | 3,383 (10.8) | 28,950 (82.9) | 2,485 (7.1) | 3,470 (9.9) | 32,862 (84.0) | 2,620 (6.7) | 3,626 (9.3) | 15,196 (84.0) | 945 (5.2) | 1,942 (10.7) |
| North West | 9,056 (75.7) | 652 (5.5) | 2,253 (18.8) | 23,698 (79.1) | 1,700 (5.7) | 4,580 (15.3) | 27,332 (80.6) | 1,803 (5.3) | 4,756 (14.0) | 31,325 (81.7) | 1,900 (5.0) | 5,099 (13.3) | 16,067 (81.0) | 860 (4.3) | 2,917 (14.7) |
| South East | 8,544 (75.8) | 690 (6.1) | 2,043 (18.1) | 21,439 (79.8) | 1,563 (5.8) | 3,862 (14.4) | 25,221 (81.8) | 1,649 (5.3) | 3,972 (12.9) | 28,888 (83.1) | 1,736 (5.0) | 4,146 (11.9) | 15,482 (81.5) | 962 (5.1) | 2,550 (13.4) |
| West Midlands | 6,636 (75.0) | 533 (6.0) | 1,676 (18.9) | 16,584 (78.7) | 1,137 (5.4) | 3,359 (15.9) | 18,883 (79.8) | 1,197 (5.1) | 3,569 (15.1) | 21,707 (81.0) | 1,286 (4.8) | 3,819 (14.2) | 11,404 (79.6) | 672 (4.7) | 2,247 (15.7) |
| South West | 5,978 (74.2) | 506 (6.3) | 1,572 (19.5) | 13,844 (78.0) | 1,085 (6.1) | 2,828 (15.9) | 15,344 (78.7) | 1,176 (6.0) | 2,974 (15.3) | 15,834 (79.6) | 1,185 (6.0) | 2,883 (14.5) | 8,632 (78.5) | 645 (5.9) | 1,725 (15.7) |
| East of England | 1,677 (75.1) | 135 (6.0) | 421 (18.9) | 4,231 (81.4) | 269 (5.2) | 698 (13.4) | 5,285 (83.7) | 296 (4.7) | 732 (11.6) | 6,559 (85.7) | 317 (4.1) | 775 (10.1) | 3,504 (84.5) | 174 (4.2) | 468 (11.3) |
| North East | 2,135 (77.2) | 135 (4.9) | 497 (18.0) | 5,058 (81.0) | 273 (4.4) | 917 (14.7) | 5,699 (81.6) | 287 (4.1) | 997 (14.3) | 6,243 (82.4) | 310 (4.1) | 1,023 (13.5) | 3,274 (80.6) | 178 (4.4) | 610 (15.0) |
| Yorkshire & The Humber | 1,704 (77.8) | 131 (6.0) | 355 (16.2) | 4,033 (82.2) | 267 (5.4) | 605 (12.3) | 4,508 (83.5) | 271 (5.0) | 618 (11.5) | 4,041 (85.3) | 215 (4.5) | 483 (10.2) | 2,026 (84.7) | 102 (4.3) | 264 (11.0) |
| East Midlands | 1,119 (78.6) | 67 (4.7) | 237 (16.7) | 2,737 (82.7) | 143 (4.3) | 430 (13.0) | 2,982 (84.0) | 139 (3.9) | 429 (12.1) | 2,631 (85.9) | 102 (3.3) | 330 (10.8) | 1,303 (84.6) | 53 (3.4) | 185 (12.0) |

| | 2019/20 | | | 2020/21 | | | 2021/22 | | | 2022/23 | | | 2023/24 | | |
|---|---|---|---|---|---|---|---|---|---|---|---|---|---|---|---|
| | **Low** 45,810 (76.3)[1] | **Moderate** 3,544 (5.9)[1] | **Severe** 10,659 (17.8)[1] | **Low** 117,328 (79.9)[1] | **Moderate** 8,786 (6.0)[1] | **Severe** 20,662 (14.1)[1] | **Low** 134,204 (81.3)[1] | **Moderate** 9,303 (5.6)[1] | **Severe** 21,517 (13.0)[1] | **Low** 150,090 (82.5)[1] | **Moderate** 9,671 (5.32)[1] | **Severe** 22,184 (12.2)[1] | **Low** 76,888 (81.5)[1] | **Moderate** 4,591 (4.9)[1] | **Severe** 12,908 (13.7)[1] |
| **Socioeconomic status**[4] | | | | | | | | | | | | | | | |
| 1 (Least deprived) | 9,182 (76.3) | 722 (6.0) | 2,132 (17.7) | 21,717 (81.4) | 1,439 (5.4) | 3,510 (13.2) | 25,053 (83.0) | 1,506 (5.0) | 3,643 (12.1) | 27,936 (84.0) | 1,551 (4.7) | 3,776 (11.4) | 15,631 (82.2) | 903 (4.7) | 2,490 (13.1) |
| 2 | 9,573 (76.9) | 739 (5.9) | 2,135 (17.2) | 22,548 (81.1) | 1,493 (5.4) | 3,768 (13.5) | 25,709 (82.5) | 1,554 (5.0) | 3,918 (12.6) | 28,812 (83.5) | 1,661 (4.8) | 4,036 (11.7) | 15,983 (81.6) | 937 (4.8) | 2,677 (13.7) |
| 3 | 9,164 (76.8) | 688 (5.8) | 2,081 (17.4) | 22,660 (80.4) | 1,590 (5.6) | 3,930 (13.9) | 25,929 (81.7) | 1,695 (5.3) | 4,117 (13.0) | 29,075 (83.0) | 1,762 (5.0) | 4,182 (11.9) | 15,181 (81.8) | 896 (4.8) | 2,473 (13.3) |
| 4 | 9,451 (76.5) | 735 (5.9) | 2,167 (17.5) | 25,420 (79.6) | 2,075 (6.5) | 4,452 (13.9) | 29,103 (80.9) | 2,227 (6.2) | 4,662 (13.0) | 32,564 (82.1) | 2,291 (5.8) | 4,822 (12.2) | 15,992 (81.6) | 958 (4.9) | 2,658 (13.6) |
| 5 (Most deprived) | 8,440 (75.1) | 660 (5.9) | 2,144 (19.1) | 24,983 (77.6) | 2,189 (6.8) | 5,002 (15.5) | 28,410 (79.1) | 2,321 (6.5) | 5,177 (14.4) | 31,703 (80.3) | 2,406 (6.1) | 5,368 (13.6) | 14,101 (80.1) | 897 (5.1) | 2,610 (14.8) |
| **Has another clinical risk comorbidity** | | | | | | | | | | | | | | | |
| No | 12,556 (78.5) | 961 (6.0) | 2,487 (15.5) | 43,455 (81.5) | 3,599 (6.7) | 6,266 (11.8) | 49,289 (82.9) | 3,760 (6.3) | 6,420 (10.8) | 54,692 (83.9) | 3,913 (6.0) | 6,570 (10.1) | 20,753 (83.6) | 1,249 (5.0) | 2,837 (11.4) |
| Yes | 33,254 (75.6) | 2,583 (5.9) | 8,172 (18.6) | 73,873 (79.0) | 5,187 (5.6) | 14,396 (15.4) | 84,915 (80.4) | 5,543 (5.3) | 15,097 (14.3) | 95,398 (81.7) | 5,758 (4.9) | 15,614 (13.4) | 56,135 (80.7) | 3,342 (4.8) | 10,071 (14.5) |

[1]n (row %)

[2]Number suppressed so that number of individuals with indetermined sex cannot be calculated.

[3]Number and proportions suppressed to avoid statistical disclosure.

[4]Measured by Index of Multiple Deprivation.

In 2019/20 and 2023/24 individuals aged ≥65 years were eligible for influenza vaccination (irrespective of additional clinical risk comorbidities). In 2020/21, 2021/22, and 2022/23 age eligibility was widened to ≥50 years due to the COVID-19 pandemic.

**Appendix 7: Number and proportion of individuals who were influenza vaccinated in each season, amongst individuals who were not additionally age eligible for influenza vaccination.**

| | 2019/20 | | 2020/21 | | 2021/22 | | 2022/23 | | 2023/24 | |
|---|---|---|---|---|---|---|---|---|---|---|
| | **Overall** 122,161 | **Influenza vaccinated** 49,513 (40.5)[1] | **Overall** 56,890 | **Influenza vaccinated** 21,014 (36.9)[1] | **Overall** 63,156 | **Influenza vaccinated** 21,460 (34.0)[1] | **Overall** 69,789[1] | **Influenza vaccinated** 21,565 (30.9)[1] | **Overall** 183,083 | **Influenza vaccinated** 73,427 (40.1)[1] |
| **Age group (years)** | | | | | | | | | | |
| <40 | 21,277 | 5,577 (26.2) | 23,821 | 7,492 (31.5) | 26,573 | 7,521 (28.3) | 29,707 | 7,431 (25.0) | 32,032 | 7,198 (22.5) |
| 40-49 | 29,927 | 9,878 (33.0) | 33,069 | 13,522 (40.9) | 36,583 | 13,939 (38.1) | 40,082 | 14,134 (35.3) | 43,936 | 13,504 (30.7) |
| 50-64 | 70,957 | 34,058 (48.0) | — | — | — | — | — | — | 107,115 | 52,725 (49.2) |
| 65-69 | — | — | — | — | — | — | — | — | — | — |
| ≥70 | — | — | — | — | — | — | — | — | — | — |
| **Sex** | | | | | | | | | | |
| Male | 69,196 | 25,783 (37.3) | 34,508 | 11,439 (33.1) | 37,628 | 11,151 (29.6) | 41,191 | 11,253 (27.3) | 100,165 | 36,936 (36.9) |
| Female[2] | 52,963 | — (44.8) | 22,382 | — (42.8) | 25,528 | — (40.4) | 28,597 | — (36.1) | 82,915 | — (44.0) |
| Indetermined[3] | <10 | <10 | <10 | <10 | <10 | <10 | <10 | <10 | <10 | <10 |
| **Ethnicity** | | | | | | | | | | |
| White | 77,511 | 32,317 (41.7) | 32,325 | 12,268 (38.0) | 37,603 | 13,585 (36.1) | 43,552 | 14,184 (32.6) | 130,911 | 56,061 (42.8) |
| Asian | 15,904 | 6,750 (42.4) | 10,318 | 4,028 (39.0) | 11,575 | 3,892 (33.6) | 13,113 | 4,130 (31.5) | 25,863 | 9,544 (36.9) |
| Black | 5,965 | 2,311 (38.7) | 3,178 | 1,077 (33.9) | 3,499 | 955 (27.3) | 3,899 | 996 (25.5) | 9,430 | 2,845 (30.2) |
| Other | 3,560 | 1,256 (35.3) | 2,330 | 859 (36.9) | 2,552 | 809 (31.7) | 2,960 | 889 (30.0) | 6,060 | 2,041 (33.7) |
| Mixed | 1,851 | 650 (35.1) | 1,076 | 362 (33.6) | 1,209 | 334 (27.6) | 1,378 | 321 (23.3) | 3,033 | 937 (30.9) |
| Missing | 17,370 | 6,229 (35.9) | 7,663 | 2,420 (31.6) | 6,718 | 1,885 (28.1) | 4,887 | 1,045 (21.4) | 7,786 | 1,999 (25.7) |
| **Region** | | | | | | | | | | |
| London | 32,017 | 12,277 (38.3) | 16,456 | 5,807 (35.3) | 17,841 | 4,978 (27.9) | 19,755 | 5,019 (25.4) | 46,818 | 15,766 (33.7) |
| North West | 23,670 | 10,096 (42.7) | 10,874 | 3,964 (36.5) | 12,251 | 4,375 (35.7) | 13,757 | 4,433 (32.2) | 37,947 | 15,902 (41.9) |
| South East | 20,776 | 8,162 (39.3) | 9,534 | 3,558 (37.3) | 10,922 | 3,830 (35.1) | 12,343 | 4,003 (32.4) | 33,577 | 14,274 (42.5) |
| West Midlands | 17,175 | 7,078 (41.2) | 7,989 | 2,966 (37.1) | 8,825 | 3,155 (35.8) | 10,121 | 3,295 (32.6) | 26,657 | 11,019 (41.3) |
| South West | 13,383 | 5,523 (41.3) | 5,656 | 2,223 (39.3) | 6,122 | 2,418 (39.5) | 6,271 | 2,258 (36.0) | 17,028 | 7,486 (44.0) |
| East of England | 3,780 | 1,472 (38.9) | 1,657 | 659 (39.8) | 2,087 | 711 (34.1) | 2,503 | 799 (31.9) | 7,507 | 3,048 (40.6) |
| North East | 4,688 | 2,143 (45.7) | 1,904 | 779 (40.9) | 2,083 | 857 (41.1) | 2,253 | 838 (37.2) | 6,287 | 2,965 (47.2) |

| | 2019/20 | | 2020/21 | | 2021/22 | | 2022/23 | | 2023/24 | |
|---|---|---|---|---|---|---|---|---|---|---|
| | **Overall** 122,161 | **Influenza vaccinated** 49,513 (40.5)[1] | **Overall** 56,890 | **Influenza vaccinated** 21,014 (36.9)[1] | **Overall** 63,156 | **Influenza vaccinated** 21,460 (34.0)[1] | **Overall** 69,789[1] | **Influenza vaccinated** 21,565 (30.9)[1] | **Overall** 183,083 | **Influenza vaccinated** 73,427 (40.1)[1] |
| Yorkshire & The Humber | 3,784 | 1,550 (41.0) | 1,647 | 630 (38.3) | 1,840 | 665 (36.1) | 1,668 | 546 (32.7) | 4,362 | 1,814 (41.6) |
| East Midlands | 2,888 | 1,212 (42.0) | 1,173 | 428 (36.5) | 1,185 | 471 (39.7) | 1,118 | 374 (33.5) | 2,900 | 1,153 (39.8) |
| **Socioeconomic status[4]** | | | | | | | | | | |
| 1 (Least deprived) | 18,499 | 7,123 (38.5) | 7,447 | 2,893 (38.8) | 8,356 | 3,162 (37.8) | 9,276 | 3,263 (35.2) | 28,009 | 12,409 (44.3) |
| 2 | 20,198 | 7,985 (39.5) | 8,550 | 3,170 (37.1) | 9,501 | 3,434 (36.1) | 10,507 | 3,514 (33.4) | 30,363 | 12,936 (42.6) |
| 3 | 22,871 | 9,031 (39.5) | 10,488 | 3,850 (36.7) | 11,545 | 3,886 (33.7) | 12,749 | 3,890 (30.5) | 34,248 | 13,747 (40.1) |
| 4 | 28,908 | 11,768 (40.7) | 14,286 | 5,225 (36.6) | 15,867 | 5,176 (32.6) | 17,481 | 5,107 (29.2) | 43,431 | 16,629 (38.3) |
| 5 (Most deprived) | 31,685 | 13,606 (42.9) | 16,119 | 5,876 (36.5) | 17,887 | 5,802 (32.4) | 19,776 | 5,791 (29.3) | 47,032 | 17,706 (37.6) |
| **Has another clinical risk comorbidity** | | | | | | | | | | |
| No | 63,065 | 13,384 (21.2) | 35,334 | 8,496 (24.0) | 38,864 | 8,433 (21.7) | 42,918 | 8,228 (19.2) | 92,688 | 22,135 (23.9) |
| Yes | 59,096 | 36,129 (61.1) | 21,556 | 12,518 (58.1) | 24,292 | 13,027 (53.6) | 26,871 | 13,337 (49.6) | 90,395 | 51,292 (56.7) |
| **Chronic liver disease severity** | | | | | | | | | | |
| Low | 98,453 | 38,095 (38.7) | 46,413 | 16,429 (35.4) | 52,625 | 17,318 (32.9) | 59,137 | 17,650 (29.8) | 157,058 | 61,570 (39.2) |
| Moderate | 11,170 | 4,702 (42.1) | 6,854 | 2,700 (39.4) | 6,914 | 2,408 (34.8) | 7,019 | 2,293 (32.7) | 12,328 | 4,726 (38.3) |
| Severe | 12,538 | 6,716 (53.6) | 3,623 | 1,885 (52.0) | 3,617 | 1,734 (47.9) | 3,633 | 1,622 (44.6) | 13,697 | 7,131 (52.1) |
| **Alcohol-related aetiology** | | | | | | | | | | |
| Not alcohol-related | 56,740 | 20,872 (36.8) | 27,736 | 8,592 (31.0) | 30,103 | 9,150 (30.4) | 32,355 | 9,035 (27.9) | 78,081 | 29,198 (37.4) |
| Uncertain if alcohol-related | 55,215 | 23,708 (42.9) | 25,880 | 11,063 (42.7) | 29,717 | 11,060 (37.2) | 34,025 | 11,354 (33.4) | 93,055 | 39,057 (42.0) |
| Alcohol-related | 10,206 | 4,933 (48.3) | 3,274 | 1,359 (41.5) | 3,336 | 1,250 (37.5) | 3,409 | 1,176 (34.5) | 11,947 | 5,172 (43.3) |
| **Viral-related aetiology** | | | | | | | | | | |
| Not viral-related | 103,445 | 40,378 (39.0) | 48,453 | 17,277 (35.7) | 54,469 | 18,068 (33.2) | 60,810 | 18,268 (30.0) | 160,727 | 63,114 (39.3) |
| Uncertain if viral-related | 14,816 | 7,629 (51.5) | 5,195 | 2,495 (48.0) | 5,337 | 2,332 (43.7) | 5,452 | 2,244 (41.2) | 17,194 | 8,658 (50.4) |

| | 2019/20 | | 2020/21 | | 2021/22 | | 2022/23 | | 2023/24 | |
|---|---|---|---|---|---|---|---|---|---|---|
| | **Overall** 122,161 | **Influenza vaccinated** 49,513 (40.5)[1] | **Overall** 56,890 | **Influenza vaccinated** 21,014 (36.9)[1] | **Overall** 63,156 | **Influenza vaccinated** 21,460 (34.0)[1] | **Overall** 69,789[1] | **Influenza vaccinated** 21,565 (30.9)[1] | **Overall** 183,083 | **Influenza vaccinated** 73,427 (40.1)[1] |
| Viral-related | 3,900 | 1,506 (38.6) | 3,242 | 1,242 (38.3) | 3,350 | 1,060 (31.6) | 3,527 | 1,053 (29.9) | 5,162 | 1,655 (32.1) |
| **Green Book listed diagnosis[5]** | | | | | | | | | | |
| No | 103,389 | 40,489 (39.2) | 48,313 | 17,338 (35.9) | 54,524 | 18,177 (33.3) | 60,980 | 18,449 (30.3) | 162,161 | 64,112 (39.5) |
| Yes | 18,772 | 9,024 (48.1) | 8,577 | 3,676 (42.9) | 8,632 | 3,283 (38.0) | 8,809 | 3,116 (35.4) | 20,922 | 9,315 (44.5) |

[1]n (row %)

[2]Number suppressed so that number of individuals with indetermined sex cannot be calculated.

[3]Number and proportions suppressed to avoid statistical disclosure.

[4]Measured by Index of Multiple Deprivation.

[5]Of: cirrhosis, biliary atresia, chronic hepatitis.

In 2019/20 and 2023/24 individuals aged ≥65 years were eligible for influenza vaccination (irrespective of additional clinical risk comorbidities). In 2020/21, 2021/22, and 2022/23 age eligibility was widened to ≥50 years due to the COVID-19 pandemic.

**Appendix 8: Number and proportion of individuals who were influenza vaccinated in each season, amongst individuals who were additionally age eligible for influenza vaccination.**

| | 2019/20 | | 2020/21 | | 2021/22 | | 2022/23 | | 2023/24 | |
|---|---|---|---|---|---|---|---|---|---|---|
| | **Overall** 60,013 | **Influenza vaccinated** 47,102 (78.5)[1] | **Overall** 146,776 | **Influenza vaccinated** 107,869 (73.5)[1] | **Overall** 165,024 | **Influenza vaccinated** 122,312 (74.1)[1] | **Overall** 181,945[1] | **Influenza vaccinated** 129,376 (71.1)[1] | **Overall** 94,387 | **Influenza vaccinated** 75,241 (79.7)[1] |
| **Age group (years)** | | | | | | | | | | |
| <40 | – | – | – | – | – | – | – | – | – | – |
| 40-49 | – | – | – | – | – | – | – | – | – | – |
| 50-64 | – | – | 79,377 | 51,205 (64.5) | 89,082 | 58,365 (65.5) | 97,786 | 60,302 (61.7) | – | – |
| 65-69 | 20,561 | 15,221 (74.0) | 22,658 | 18,417 (81.3) | 25,449 | 20,612 (81.0) | 28,079 | 21,979 (78.3) | 31,525 | 23,658 (75.0) |
| ≥70 | 39,452 | 31,881 (80.8) | 44,741 | 38,247 (85.5) | 50,493 | 43,335 (85.8) | 56,080 | 47,095 (84.0) | 62,862 | 51,583 (82.1) |
| **Sex** | | | | | | | | | | |
| Male | 27,764 | 21,857 (78.7) | 73,665 | 53,119 (72.1) | 82,313 | 60,104 (73.0) | 90,406 | 63,439 (70.2) | 43,970 | 35,492 (80.7) |
| Female[2] | 32,249 | – (78.3) | 73,109 | – (74.9) | 82,709 | – (75.2) | 91,538 | – (72.0) | 50,417 | – (78.8) |
| Indetermined[3] | <10 | <10 | <10 | <10 | <10 | <10 | <10 | <10 | <10 | <10 |
| **Ethnicity** | | | | | | | | | | |
| White | 45,087 | 35,722 (79.2) | 105,303 | 79,442 (75.4) | 125,434 | 96,212 (76.7) | 144,605 | 107,010 (74.0) | 80,266 | 65,657 (81.8) |
| Asian | 3,527 | 2,807 (79.6) | 11,914 | 8,431 (70.8) | 13,552 | 9,197 (67.9) | 15,857 | 10,055 (63.4) | 6,631 | 4,715 (71.1) |
| Black | 1,383 | 959 (69.3) | 5,076 | 2,730 (53.8) | 5,884 | 2,947 (50.1) | 6,780 | 3,295 (48.6) | 2,516 | 1,375 (54.7) |
| Other | 646 | 486 (75.2) | 2,561 | 1,656 (64.7) | 3,042 | 1,926 (63.3) | 3,634 | 2,169 (59.7) | 1,498 | 1,074 (71.7) |
| Mixed | 319 | 225 (70.5) | 1,369 | 840 (61.4) | 1,579 | 951 (60.2) | 1,902 | 1,099 (57.8) | 690 | 460 (66.7) |
| Missing | 9,051 | 6,903 (76.3) | 20,553 | 14,770 (71.9) | 15,533 | 11,079 (71.3) | 9,167 | 5,748 (62.7) | 2,786 | 1,960 (70.4) |
| **Region** | | | | | | | | | | |
| London | 11,261 | 8,437 (74.9) | 31,436 | 21,019 (66.9) | 34,905 | 22,017 (63.1) | 39,108 | 23,873 (61.0) | 18,083 | 12,834 (71.0) |
| North West | 11,961 | 9,567 (80.0) | 29,978 | 21,987 (73.3) | 33,891 | 25,432 (75.0) | 38,324 | 27,353 (71.4) | 19,844 | 16,031 (80.8) |
| South East | 11,277 | 8,935 (79.2) | 26,864 | 20,266 (75.4) | 30,842 | 23,975 (77.7) | 34,770 | 26,194 (75.3) | 18,994 | 15,672 (82.5) |
| West Midlands | 8,845 | 6,907 (78.1) | 21,080 | 15,770 (74.8) | 23,649 | 17,906 (75.7) | 26,812 | 19,230 (71.7) | 14,323 | 11,499 (80.3) |
| South West | 8,056 | 6,408 (79.5) | 17,757 | 13,652 (76.9) | 19,494 | 15,487 (79.4) | 19,902 | 15,315 (77.0) | 11,002 | 9,096 (82.7) |
| East of England | 2,233 | 1,746 (78.2) | 5,198 | 4,034 (77.6) | 6,313 | 5,027 (79.6) | 7,651 | 5,772 (75.4) | 4,146 | 3,533 (85.2) |
| North East | 2,767 | 2,239 (80.9) | 6,248 | 4,903 (78.5) | 6,983 | 5,583 (80.0) | 7,576 | 5,808 (76.7) | 4,062 | 3,367 (82.9) |

| | 2019/20 | | 2020/21 | | 2021/22 | | 2022/23 | | 2023/24 | |
|---|---|---|---|---|---|---|---|---|---|---|
| | **Overall** 60,013 | **Influenza vaccinated** 47,102 (78.5)[1] | **Overall** 146,776 | **Influenza vaccinated** 107,869 (73.5)[1] | **Overall** 165,024 | **Influenza vaccinated** 122,312 (74.1)[1] | **Overall** 181,945[1] | **Influenza vaccinated** 129,376 (71.1)[1] | **Overall** 94,387 | **Influenza vaccinated** 75,241 (79.7)[1] |
| Yorkshire & The Humber | 2,190 | 1,760 (80.4) | 4,905 | 3,745 (76.4) | 5,397 | 4,104 (76.0) | 4,739 | 3,580 (75.5) | 2,392 | 1,984 (82.9) |
| East Midlands | 1,423 | 1,103 (77.5) | 3,310 | 2,493 (75.3) | 3,550 | 2,781 (78.3) | 3,063 | 2,251 (73.5) | 1,541 | 1,225 (79.5) |
| **Socioeconomic status[4]** | | | | | | | | | | |
| 1 (Least deprived) | 12,036 | 9,832 (81.7) | 26,666 | 20,969 (78.6) | 30,202 | 24,572 (81.4) | 33,263 | 26,353 (79.2) | 19,024 | 16,351 (85.9) |
| 2 | 12,447 | 9,948 (79.9) | 27,809 | 21,440 (77.1) | 31,181 | 24,678 (79.1) | 34,509 | 26,351 (76.4) | 19,597 | 16,346 (83.4) |
| 3 | 11,933 | 9,331 (78.2) | 28,180 | 20,792 (73.8) | 31,741 | 23,735 (74.8) | 35,019 | 25,086 (71.6) | 18,550 | 14,831 (80.0) |
| 4 | 12,353 | 9,444 (76.5) | 31,947 | 22,616 (70.8) | 35,992 | 25,210 (70.0) | 39,677 | 26,435 (66.6) | 19,608 | 14,904 (76.0) |
| 5 (Most deprived) | 11,244 | 8,547 (76.0) | 32,174 | 22,052 (68.5) | 35,908 | 24,117 (67.2) | 39,477 | 25,151 (63.7) | 17,608 | 12,809 (72.7) |
| **Has another clinical risk comorbidity** | | | | | | | | | | |
| No | 16,004 | 11,457 (71.6) | 53,320 | 33,013 (61.9) | 59,469 | 38,961 (65.5) | 65,175 | 39,929 (61.3) | 24,839 | 19,181 (77.2) |
| Yes | 44,009 | 35,645 (81.0) | 93,456 | 74,856 (80.1) | 105,555 | 83,351 (79.0) | 116,770 | 89,447 (76.6) | 69,548 | 56,060 (80.6) |
| **Chronic liver disease severity** | | | | | | | | | | |
| Low | 45,810 | 36,345 (79.3) | 117,328 | 86,467 (73.7) | 134,204 | 100,042 (74.5) | 150,090 | 107,064 (71.3) | 76,888 | 61,715 (80.3) |
| Moderate | 3,544 | 2,725 (76.9) | 8,786 | 6,102 (69.5) | 9,303 | 6,356 (68.3) | 9,671 | 6,446 (66.7) | 4,591 | 3,554 (77.4) |
| Severe | 10,659 | 8,032 (75.4) | 20,662 | 15,300 (74.0) | 21,517 | 15,914 (74.0) | 22,184 | 15,866 (71.5) | 12,908 | 9,972 (77.3) |
| **Alcohol-related aetiology** | | | | | | | | | | |
| Not alcohol-related | 26,068 | 20,789 (79.7) | 63,552 | 46,724 (73.5) | 69,940 | 52,273 (74.7) | 75,316 | 54,106 (71.8) | 37,972 | 30,615 (80.6) |
| Uncertain if alcohol-related | 28,856 | 22,603 (78.3) | 70,222 | 52,049 (74.1) | 81,256 | 60,395 (74.3) | 92,177 | 65,631 (71.2) | 49,792 | 39,604 (79.5) |
| Alcohol-related | 5,089 | 3,710 (72.9) | 13,002 | 9,096 (70.0) | 13,828 | 9,644 (69.7) | 14,452 | 9,639 (66.7) | 6,623 | 5,022 (75.8) |
| **Viral-related aetiology** | | | | | | | | | | |
| Not viral-related | 48,455 | 38,286 (79.0) | 122,873 | 90,317 (73.5) | 139,506 | 103,679 (74.3) | 154,959 | 110,266 (71.2) | 79,133 | 63,427 (80.2) |
| Uncertain if viral-related | 11,247 | 8,591 (76.4) | 22,308 | 16,564 (74.3) | 23,684 | 17,577 (74.2) | 24,910 | 17,960 (72.1) | 14,634 | 11,402 (77.9) |

| | 2019/20 | | 2020/21 | | 2021/22 | | 2022/23 | | 2023/24 | |
|---|---|---|---|---|---|---|---|---|---|---|
| | **Overall** 60,013 | **Influenza vaccinated** 47,102 (78.5)[1] | **Overall** 146,776 | **Influenza vaccinated** 107,869 (73.5)[1] | **Overall** 165,024 | **Influenza vaccinated** 122,312 (74.1)[1] | **Overall** 181,945[1] | **Influenza vaccinated** 129,376 (71.1)[1] | **Overall** 94,387 | **Influenza vaccinated** 75,241 (79.7)[1] |
| Viral-related | 311 | 225 (72.3) | 1,595 | 988 (61.9) | 1,834 | 1,056 (57.6) | 2,076 | 1,150 (55.4) | 620 | 412 (66.5) |
| **Green Book listed diagnosis[5]** | | | | | | | | | | |
| No | 50,037 | 39,604 (79.1) | 124,802 | 92,055 (73.8) | 141,920 | 105,913 (74.6) | 157,885 | 112,814 (71.5) | 81,594 | 65,498 (80.3) |
| Yes | 9,976 | 7,498 (75.2) | 21,974 | 15,814 (72.0) | 23,104 | 16,399 (71.0) | 24,060 | 16,562 (68.8) | 12,793 | 9,743 (76.2) |

[1]n (row %)

[2]Number suppressed so that number of individuals with indetermined sex cannot be calculated.

[3]Number and proportions suppressed to avoid statistical disclosure.

[4]Measured by Index of Multiple Deprivation.

[5]Of: cirrhosis, biliary atresia, chronic hepatitis.

In 2019/20 and 2023/24 individuals aged ≥65 years were eligible for influenza vaccination (irrespective of additional clinical risk comorbidities). In 2020/21, 2021/22, and 2022/23 age eligibility was widened to ≥50 years due to the COVID-19 pandemic.

**Appendix 9: Incidence rate ratios of influenza vaccine uptake by severity of chronic liver disease and age eligibility for influenza vaccination.**

| | Eligible due to chronic liver disease (not additionally age eligible) | | | | | Eligible due to chronic liver disease and age | | | | |
|---|---|---|---|---|---|---|---|---|---|---|
| **Severity** | **Model 1** | **Model 2** | **Model 3** | **Model 4** | **Sensitivity** | **Model 1** | **Model 2** | **Model 3** | **Model 4** | **Sensitivity** |
| **2019/20** | | | | | | | | | | |
| Low | Ref | Ref | Ref | Ref | Ref | Ref | Ref | Ref | Ref | Ref |
| Moderate | 1.28 (1.24–1.32) | 1.40 (1.35–1.44) | 1.40 (1.36–1.45) | 1.45 (1.40–1.50) | 1.42 (1.37–1.48) | 0.93 (0.89–0.98) | 0.94 (0.90–0.98) | 0.94 (0.90–0.98) | 0.94 (0.90–0.99) | 0.94 (0.89–0.99) |
| Severe | 1.51 (1.47–1.55) | 1.31 (1.27–1.35) | 1.32 (1.28–1.35) | 1.35 (1.30–1.39) | 1.33 (1.29–1.37) | 0.90 (0.87–0.92) | 0.89 (0.86–0.91) | 0.88 (0.86–0.91) | 0.90 (0.87–0.92) | 0.89 (0.87–0.92) |
| **2020/21** | | | | | | | | | | |
| Low | Ref | Ref | Ref | Ref | Ref | Ref | Ref | Ref | Ref | Ref |
| Moderate | 1.21 (1.16–1.26) | 1.39 (1.33–1.45) | 1.39 (1.32–1.45) | 1.42 (1.35–1.49) | 1.37 (1.31–1.44) | 0.91 (0.88–0.94) | 0.92 (0.90–0.95) | 0.95 (0.92–0.98) | 0.97 (0.93–1.00) | 0.94 (0.91–0.97) |
| Severe | 1.74 (1.65–1.83) | 1.46 (1.38–1.54) | 1.46 (1.39–1.55) | 1.52 (1.43–1.61) | 1.49 (1.40–1.58) | 0.97 (0.95–0.99) | 0.94 (0.92–0.96) | 0.94 (0.93–0.96) | 0.95 (0.93–0.97) | 0.95 (0.93–0.97) |
| **2021/22** | | | | | | | | | | |
| Low | Ref | Ref | Ref | Ref | Ref | Ref | Ref | Ref | Ref | Ref |
| Moderate | 1.11 (1.06–1.16) | 1.29 (1.23–1.35) | 1.31 (1.25–1.38) | 1.36 (1.29–1.43) | 1.31 (1.24–1.37) | 0.84 (0.82–0.87) | 0.85 (0.83–0.88) | 0.89 (0.86–0.91) | 0.92 (0.89–0.95) | 0.89 (0.87–0.92) |
| Severe | 1.67 (1.58–1.76) | 1.43 (1.35–1.51) | 1.41 (1.34–1.50) | 1.45 (1.37–1.54) | 1.44 (1.36–1.52) | 0.93 (0.91–0.95) | 0.92 (0.90–0.93) | 0.91 (0.89–0.93) | 0.91 (0.90–0.93) | 0.92 (0.90–0.94) |
| **2022/23** | | | | | | | | | | |
| Low | Ref | Ref | Ref | Ref | Ref | Ref | Ref | Ref | Ref | Ref |
| Moderate | 1.16 (1.11–1.21) | 1.35 (1.28–1.41) | 1.37 (1.31–1.44) | 1.40 (1.33–1.47) | 1.35 (1.29–1.42) | 0.88 (0.85–0.90) | 0.89 (0.87–0.92) | 0.92 (0.90–0.95) | 0.95 (0.93–0.98) | 0.93 (0.90–0.96) |
| Severe | 1.70 (1.61–1.80) | 1.43 (1.35–1.52) | 1.43 (1.35–1.52) | 1.45 (1.37–1.54) | 1.43 (1.35–1.52) | 0.96 (0.94–0.98) | 0.94 (0.93–0.96) | 0.94 (0.92–0.96) | 0.94 (0.92–0.95) | 0.94 (0.93–0.96) |
| **2023/24** | | | | | | | | | | |
| Low | Ref | Ref | Ref | Ref | Ref | Ref | Ref | Ref | Ref | Ref |
| Moderate | 1.09 (1.05–1.12) | 1.18 (1.14–1.22) | 1.24 (1.20–1.28) | 1.27 (1.22–1.32) | 1.23 (1.18–1.28) | 0.90 (0.87–0.94) | 0.91 (0.87–0.95) | 0.92 (0.88–0.95) | 0.94 (0.90–0.98) | 0.92 (0.88–0.97) |
| Severe | 1.40 (1.36–1.44) | 1.24 (1.21–1.28) | 1.25 (1.22–1.29) | 1.22 (1.18–1.27) | 1.23 (1.19–1.27) | 0.91 (0.89–0.94) | 0.91 (0.89–0.93) | 0.89 (0.87–0.92) | 0.89 (0.86–0.91) | 0.90 (0.87–0.92) |

Incidence rate ratios (95% confidence intervals) calculated using Poisson regression with robust standard errors.

Model 1 adjusted for: age group (all individuals).

Model 2 adjusted for: age group, other clinical risk comorbidity (all individuals).

Model 3 adjusted for: age group, other clinical risk comorbidity, sex, socioeconomic status, geographical region (all individuals).

Model 4 (fully) adjusted for: age group, other clinical risk comorbidity, sex, socioeconomic status, geographical region, ethnicity (thus restricted to those with complete ethnicity).

Sensitivity: model 3 but limited to those with complete ethnicity.

In 2019/20 and 2023/24 individuals aged ≥65 years were eligible for influenza vaccination (irrespective of additional clinical risk comorbidities). In 2020/21, 2021/22, and 2022/23 age eligibility was widened to ≥50 years due to the COVID-19 pandemic.

**Appendix 10: Number and proportion vaccinated and incidence rate ratios of influenza vaccine uptake by severity of chronic liver disease, stratified by clinical risk comorbidity and age eligibility for influenza vaccination (2023/24 cohort).**

| | Eligible due to chronic liver disease (not additionally age eligible) | | | | Eligible due to chronic liver disease and age | | | |
|---|---|---|---|---|---|---|---|---|
| | No additional clinical risk comorbidity | | Additional clinical risk comorbidity | | No additional clinical risk comorbidity | | Additional clinical risk comorbidity | |
| **Chronic liver disease severity** | **Influenza vaccinated** 22,135 (23.9%) | **IRR (95% CI)** | **Influenza vaccinated** 51,291 (56.7%) | **IRR (95% CI)** | **Influenza vaccinated** 19,181 (77.2%) | **IRR (95% CI)** | **Influenza vaccinated** 56,060 (80.6%) | **IRR (95% CI)** |
| **2023/24** | | | | | | | | |
| Low | 17,407 (22.0) | Ref | 44,162 (56.7) | Ref | 16,211 (78.1) | Ref | 45,504 (81.1) | Ref |
| Moderate | 2,409 (30.3) | 1.89 (1.80–1.99) | 2,317 (52.8) | 0.99 (0.94–1.04) | 904 (72.4) | 0.85 (0.78–0.93) | 2,650 (79.3) | 0.97 (0.92–1.02) |
| Severe | 2,319 (41.9) | 2.12 (2.01–2.23) | 4,812 (58.9) | 1.02 (0.98–1.06) | 2,066 (72.8) | 0.80 (0.76–0.85) | 87,906 (78.5) | 0.91 (0.89–0.94) |

Poisson regression with robust standard errors. Fully adjusted (model 4) used (adjusted for age group, sex, socioeconomic status, geographical region, and ethnicity).

In 2019/20 and 2023/24 individuals aged ≥65 years were eligible for influenza vaccination (irrespective of additional clinical risk comorbidities). In 2020/21, 2021/22, and 2022/23 age eligibility was widened to ≥50 years due to the COVID-19 pandemic.

IRR = incidence rate ratios; CI = confidence intervals.

**Appendix 11: Incidence rate ratios of influenza vaccine uptake by severity of chronic liver disease, stratified by ethnicity and age eligibility for influenza vaccination (2023/24 cohort).**

| | Eligible due to chronic liver disease (not additionally age eligible) | | | | | Eligible due to chronic liver disease and age | | | | |
|---|---|---|---|---|---|---|---|---|---|---|
| **Chronic liver disease severity** | **White** | **Black** | **Asian** | **Mixed** | **Other** | **White** | **Black** | **Asian** | **Mixed** | **Other** |
| **2023/24** | | | | | | | | | | |
| Low | Ref | Ref | Ref | Ref | Ref | Ref | Ref | Ref | Ref | Ref |
| Moderate | 1.12 (1.07–1.17) | 1.49 (1.34–1.64) | 1.61 (1.46–1.78) | 1.49 (1.21–1.84) | 2.10 (1.86–2.38) | 0.92 (0.87–0.96) | 0.91 (0.73–1.14) | 1.16 (0.98–1.37) | 0.97 (0.64–1.47) | 1.15 (0.90–1.46) |
| Severe | 1.24 (1.20–1.28) | 1.27 (1.07–1.50) | 1.33 (1.19–1.48) | 1.04 (0.75–1.44) | 1.64 (1.32–2.03) | 0.88 (0.86–0.91) | 0.85 (0.68–1.06) | 0.93 (0.83–1.06) | 0.86 (0.58–1.25) | 1.23 (0.97–1.56) |

Incidence rate ratios (95% confidence intervals) calculated using Poisson regression with robust standard errors.

Fully adjusted (model 4) used (adjusted for age group, other clinical risk comorbidity, sex, socioeconomic status, and geographical region).

In 2019/20 and 2023/24 individuals aged ≥65 years were eligible for influenza vaccination (irrespective of additional clinical risk comorbidities). In 2020/21, 2021/22, and 2022/23 age eligibility was widened to ≥50 years due to the COVID-19 pandemic.

**Appendix 12: Incidence rate ratios of influenza vaccine uptake by severity of chronic liver disease, stratified by Index of Multiple Deprivation (IMD) and age eligibility for influenza vaccination (2023/24 cohort).**

| | Eligible due to chronic liver disease (not additionally age eligible) | | | | | Eligible due to chronic liver disease and age | | | | |
|---|---|---|---|---|---|---|---|---|---|---|
| **Chronic liver disease severity** | **IMD 1 (Least deprived)** | **IMD 2** | **IMD 3** | **IMD 4** | **IMD 5 (Most deprived)** | **IMD 1 (Least deprived)** | **IMD 2** | **IMD 3** | **IMD 4** | **IMD 5 (Most deprived)** |
| **2023/24** | | | | | | | | | | |
| Low | Ref | Ref | Ref | Ref | Ref | Ref | Ref | Ref | Ref | Ref |
| Moderate | 1.64 (1.49–1.81) | 1.33 (1.22–1.46) | 1.37 (1.26–1.49) | 1.20 (1.12–1.29) | 1.09 (1.02–1.17) | 0.99 (0.91–1.08) | 0.96 (0.88–1.05) | 0.90 (0.82–0.99) | 0.95 (0.87–1.05) | 0.86 (0.78–0.96) |
| Severe | 1.43 (1.32–1.54) | 1.25 (1.16–1.35) | 1.33 (1.24–1.43) | 1.23 (1.15–1.31) | 1.13 (1.07–1.20) | 0.92 (0.87–0.98) | 0.86 (0.82–0.92) | 0.89 (0.84–0.94) | 0.88 (0.83–0.93) | 0.90 (0.84–0.95) |

Incidence rate ratios (95% confidence intervals) calculated using Poisson regression with robust standard errors.

Fully adjusted (model 4) used (adjusted for age group, sex, other clinical risk comorbidity, geographical region, and ethnicity).

In 2019/20 and 2023/24 individuals aged ≥65 years were eligible for influenza vaccination (irrespective of additional clinical risk comorbidities). In 2020/21, 2021/22, and 2022/23 age eligibility was widened to ≥50 years due to the COVID-19 pandemic.

**Appendix 13: Incidence rate ratios of influenza vaccine uptake by alcohol aetiology of chronic liver disease and age eligibility for influenza vaccination.**

| | Eligible due to chronic liver disease (not additionally age eligible) | | | | Eligible due to chronic liver disease and age | | | |
|---|---|---|---|---|---|---|---|---|
| **Aetiology** | **Model 1** | **Model 2** | **Model 3** | **Model 4** | **Model 1** | **Model 2** | **Model 3** | **Model 4** |
| **2019/20** | | | | | | | | |
| Not alcohol-related | Ref | Ref | Ref | Ref | Ref | Ref | Ref | Ref |
| Uncertain if alcohol-related | 1.20 (1.17–1.22) | 1.18 (1.15–1.20) | 1.18 (1.16–1.20) | 1.19 (1.16–1.21) | 0.97 (0.94–0.99) | 0.97 (0.94–0.99) | 0.96 (0.94–0.98) | 0.96 (0.94–0.99) |
| Alcohol-related | 1.34 (1.29–1.38) | 1.33 (1.29–1.38) | 1.37 (1.32–1.41) | 1.39 (1.34–1.45) | 0.81 (0.78–0.85) | 0.83 (0.79–0.86) | 0.81 (0.78–0.85) | 0.82 (0.78–0.86) |
| **2020/21** | | | | | | | | |
| Not alcohol-related | Ref | Ref | Ref | Ref | Ref | Ref | Ref | Ref |
| Uncertain if alcohol-related | 1.54 (1.50–1.59) | 1.47 (1.43–1.52) | 1.49 (1.44–1.53) | 1.50 (1.45–1.55) | 1.02 (1.01–1.04) | 1.02 (1.00–1.03) | 1.02 (1.00–1.03) | 1.02 (1.00–1.04) |
| Alcohol-related | 1.43 (1.34–1.52) | 1.41 (1.33–1.51) | 1.47 (1.38–1.57) | 1.51 (1.41–1.62) | 0.91 (0.89–0.94) | 0.92 (0.90–0.95) | 0.93 (0.91–0.96) | 0.93 (0.90–0.96) |
| **2021/22** | | | | | | | | |
| Not alcohol-related | Ref | Ref | Ref | Ref | Ref | Ref | Ref | Ref |
| Uncertain if alcohol-related | 1.29 (1.25–1.33) | 1.25 (1.22–1.29) | 1.26 (1.22–1.30) | 1.26 (1.23–1.30) | 0.98 (0.97–0.99) | 0.98 (0.97–0.99) | 0.98 (0.96–0.99) | 0.98 (0.97–0.99) |
| Alcohol-related | 1.24 (1.16–1.32) | 1.26 (1.18–1.35) | 1.29 (1.21–1.37) | 1.31 (1.22–1.40) | 0.87 (0.85–0.89) | 0.88 (0.86–0.90) | 0.88 (0.85–0.90) | 0.87 (0.85–0.89) |
| **2022/23** | | | | | | | | |
| Not alcohol-related | Ref | Ref | Ref | Ref | Ref | Ref | Ref | Ref |
| Uncertain if alcohol-related | 1.24 (1.21–1.28) | 1.21 (1.17–1.25) | 1.21 (1.18–1.25) | 1.22 (1.19–1.26) | 0.98 (0.97–0.99) | 0.98 (0.97–1.00) | 0.98 (0.96–0.99) | 0.97 (0.96–0.99) |
| Alcohol-related | 1.24 (1.16–1.32) | 1.28 (1.19–1.37) | 1.30 (1.22–1.39) | 1.33 (1.24–1.42) | 0.88 (0.86–0.90) | 0.90 (0.87–0.92) | 0.89 (0.87–0.91) | 0.87 (0.85–0.89) |
| **2023/24** | | | | | | | | |
| Not alcohol-related | Ref | Ref | Ref | Ref | Ref | Ref | Ref | Ref |
| Uncertain if alcohol-related | 1.15 (1.13–1.17) | 1.13 (1.11–1.15) | 1.13 (1.11–1.15) | 1.13 (1.11–1.15) | 0.97 (0.95–0.99) | 0.97 (0.95–0.99) | 0.95 (0.94–0.97) | 0.95 (0.93–0.97) |
| Alcohol-related | 1.10 (1.06–1.14) | 1.12 (1.08–1.16) | 1.13 (1.09–1.17) | 1.12 (1.08–1.16) | 0.87 (0.84–0.91) | 0.88 (0.85–0.91) | 0.85 (0.82–0.88) | 0.83 (0.80–0.86) |

Incidence rate ratios (95% confidence intervals) calculated using Poisson regression with robust standard errors.

Model 1 adjusted for: age group (all individuals).

Model 2 adjusted for: age group, other clinical risk comorbidity (all individuals).

Model 3 adjusted for: age group, other clinical risk comorbidity, sex, socioeconomic status, geographical region (all individuals).

Model 4 (fully) adjusted for: age group, other clinical risk comorbidity, sex, socioeconomic status, geographical region, ethnicity (thus restricted to those with complete ethnicity).

In 2019/20 and 2023/24 individuals aged ≥65 years were eligible for influenza vaccination (irrespective of additional clinical risk comorbidities). In 2020/21, 2021/22, and 2022/23 age eligibility was widened to ≥50 years due to the COVID-19 pandemic.

**Appendix 14: Incidence rate ratios of influenza vaccine uptake by viral aetiology of chronic liver disease and age eligibility for influenza vaccination.**

| | Eligible due to chronic liver disease (not additionally age eligible) | | | | Eligible due to chronic liver disease and age | | | |
|---|---|---|---|---|---|---|---|---|
| **Aetiology** | **Model 1** | **Model 2** | **Model 3** | **Model 4** | **Model 1** | **Model 2** | **Model 3** | **Model 4** |
| **2019/20** | | | | | | | | |
| Not viral-related | Ref | Ref | Ref | Ref | Ref | Ref | Ref | Ref |
| Uncertain if viral-related | 1.46 (1.42–1.50) | 1.33 (1.29–1.37) | 1.33 (1.30–1.37) | 1.37 (1.33–1.41) | 0.93 (0.91–0.96) | 0.93 (0.90–0.95) | 0.92 (0.90–0.95) | 0.93 (0.91–0.96) |
| Viral-related | 1.25 (1.18–1.32) | 1.57 (1.48–1.67) | 1.59 (1.50–1.69) | 1.64 (1.54–1.74) | 0.79 (0.68–0.92) | 0.79 (0.68–0.92) | 0.84 (0.72–0.98) | 0.90 (0.76–1.05) |
| **2020/21** | | | | | | | | |
| Not viral-related | Ref | Ref | Ref | Ref | Ref | Ref | Ref | Ref |
| Uncertain if viral-related | 1.53 (1.46–1.61) | 1.42 (1.35–1.49) | 1.42 (1.35–1.49) | 1.47 (1.40–1.55) | 0.99 (0.97–1.01) | 0.97 (0.95–0.99) | 0.97 (0.95–0.99) | 0.98 (0.96–1.00) |
| Viral-related | 1.17 (1.10–1.24) | 1.54 (1.44–1.64) | 1.54 (1.44–1.65) | 1.61 (1.50–1.73) | 0.82 (0.76–0.88) | 0.85 (0.79–0.92) | 0.94 (0.87–1.01) | 1.05 (0.97–1.13) |
| **2021/22** | | | | | | | | |
| Not viral-related | Ref | Ref | Ref | Ref | Ref | Ref | Ref | Ref |
| Uncertain if viral-related | 1.46 (1.40–1.53) | 1.37 (1.30–1.44) | 1.37 (1.30–1.43) | 1.41 (1.34–1.48) | 0.95 (0.94–0.97) | 0.94 (0.93–0.96) | 0.94 (0.92–0.96) | 0.95 (0.93–0.97) |
| Viral-related | 0.98 (0.92–1.04) | 1.28 (1.19–1.37) | 1.36 (1.27–1.46) | 1.45 (1.35–1.56) | 0.66 (0.62–0.71) | 0.68 (0.64–0.73) | 0.81 (0.76–0.86) | 0.92 (0.85–0.98) |
| **2022/23** | | | | | | | | |
| Not viral-related | Ref | Ref | Ref | Ref | Ref | Ref | Ref | Ref |
| Uncertain if viral-related | 1.52 (1.45–1.59) | 1.39 (1.32–1.46) | 1.40 (1.33–1.47) | 1.43 (1.36–1.51) | 0.98 (0.96–1.00) | 0.97 (0.95–0.99) | 0.97 (0.95–0.98) | 0.97 (0.95–0.99) |
| Viral-related | 1.04 (0.97–1.11) | 1.37 (1.28–1.46) | 1.46 (1.36–1.56) | 1.52 (1.41–1.63) | 0.70 (0.65–0.74) | 0.72 (0.67–0.77) | 0.84 (0.79–0.90) | 0.96 (0.90–1.03) |
| **2023/24** | | | | | | | | |
| Not viral-related | Ref | Ref | Ref | Ref | Ref | Ref | Ref | Ref |
| Uncertain if viral-related | 1.37 (1.34–1.41) | 1.26 (1.22–1.29) | 1.27 (1.24–1.31) | 1.28 (1.25–1.32) | 0.93 (0.91–0.96) | 0.93 (0.91–0.95) | 0.91 (0.89–0.94) | 0.92 (0.89–0.94) |
| Viral-related | 0.94 (0.89–1.00) | 1.11 (1.05–1.18) | 1.26 (1.19–1.33) | 1.38 (1.30–1.46) | 0.64 (0.57–0.72) | 0.64 (0.57–0.72) | 0.74 (0.66–0.84) | 0.84 (0.74–0.95) |

Incidence rate ratios (95% confidence intervals) calculated using Poisson regression with robust standard errors.

Model 1 adjusted for: age group (all individuals).

Model 2 adjusted for: age group, other clinical risk comorbidity (all individuals).

Model 3 adjusted for: age group, other clinical risk comorbidity, sex, socioeconomic status, geographical region (all individuals).

Model 4 (fully) adjusted for: age group, other clinical risk comorbidity, sex, socioeconomic status, geographical region, ethnicity (thus restricted to those with complete ethnicity).

In 2019/20 and 2023/24 individuals aged ≥65 years were eligible for influenza vaccination (irrespective of additional clinical risk comorbidities). In 2020/21, 2021/22, and 2022/23 age eligibility was widened to ≥50 years due to the COVID-19 pandemic.

**Appendix 15: Incidence rate ratios of influenza vaccine uptake by Green Book diagnosis and age eligibility for influenza vaccination.**

| | Eligible due to chronic liver disease (not additionally age eligible) | | | | Eligible due to chronic liver disease and age | | | |
|---|---|---|---|---|---|---|---|---|
| **Green Book diagnosis** | **Model 1** | **Model 2** | **Model 3** | **Model 4** | **Model 1** | **Model 2** | **Model 3** | **Model 4** |
| **2019/20** | | | | | | | | |
| No | Ref | Ref | Ref | Ref | Ref | Ref | Ref | Ref |
| Yes | 1.39 (1.36–1.42) | 1.35 (1.31–1.38) | 1.36 (1.32–1.39) | 1.40 (1.36–1.44) | 0.89 (0.87–0.92) | 0.88 (0.86–0.91) | 0.88 (0.86–0.91) | 0.90 (0.87–0.92) |
| **2020/21** | | | | | | | | |
| No | Ref | Ref | Ref | Ref | Ref | Ref | Ref | Ref |
| Yes | 1.32 (1.27–1.37) | 1.40 (1.35–1.46) | 1.41 (1.36–1.47) | 1.45 (1.39–1.51) | 0.95 (0.93–0.97) | 0.93 (0.91–0.95) | 0.95 (0.93–0.97) | 0.96 (0.94–0.98) |
| **2021/22** | | | | | | | | |
| No | Ref | Ref | Ref | Ref | Ref | Ref | Ref | Ref |
| Yes | 1.21 (1.16–1.25) | 1.29 (1.24–1.35) | 1.33 (1.28–1.38) | 1.39 (1.33–1.45) | 0.88 (0.86–0.89) | 0.87 (0.85–0.89) | 0.89 (0.87–0.91) | 0.90 (0.89–0.92) |
| **2022/23** | | | | | | | | |
| No | Ref | Ref | Ref | Ref | Ref | Ref | Ref | Ref |
| Yes | 1.24 (1.19–1.29) | 1.34 (1.29–1.40) | 1.37 (1.32–1.43) | 1.41 (1.35–1.47) | 0.91 (0.90–0.93) | 0.90 (0.89–0.92) | 0.92 (0.91–0.94) | 0.93 (0.92–0.95) |
| **2023/24** | | | | | | | | |
| No | Ref | Ref | Ref | Ref | Ref | Ref | Ref | Ref |
| Yes | 1.21 (1.18–1.24) | 1.18 (1.15–1.21) | 1.23 (1.20–1.26) | 1.26 (1.22–1.29) | 0.88 (0.86–0.90) | 0.88 (0.86–0.90) | 0.88 (0.86–0.90) | 0.88 (0.86–0.91) |

Incidence rate ratios (95% confidence intervals) calculated using Poisson regression with robust standard errors.

Model 1 adjusted for: age group (all individuals).

Model 2 adjusted for: age group, other clinical risk comorbidity (all individuals).

Model 3 adjusted for: age group, other clinical risk comorbidity, sex, socioeconomic status, geographical region (all individuals).

Model 4 (fully) adjusted for: age group, other clinical risk comorbidity, sex, socioeconomic status, geographical region, ethnicity (thus restricted to those with complete ethnicity).

In 2019/20 and 2023/24 individuals aged ≥65 years were eligible for influenza vaccination (irrespective of additional clinical risk comorbidities). In 2020/21, 2021/22, and 2022/23 age eligibility was widened to ≥50 years due to the COVID-19 pandemic.
